# The role of sex hormones in severe mental illness: a genetic exploration

**DOI:** 10.1101/2024.06.25.24309226

**Authors:** R. R. Veeneman, K. J. H. Verweij, I. E. C. Sommer, J. L. Treur, J. M. Vermeulen

## Abstract

**Introduction:** Despite advances in psychotherapies and pharmacological treatments, the burden of severe mental illness (SMI) remains high. Prominent sex differences exist in SMI, with increasing evidence pointing towards a pivotal role for sex hormones. Elucidation of these hormonal influences is crucial to tailor sex-specific prevention and treatment.

**Methods:** To investigate potential shared genetics and bi-directional causal effects between sex hormone traits and SMI, we computed genetic correlations using linkage disequilibrium score regression and bi-directional summary-level Mendelian Randomization (MR). A range of sensitivity methods was applied and potential mediators were investigated using multivariable MR. Sex-stratified data from genome-wide association studies (GWAS)s were used, further stratified on menopausal status when available. The main analysis focused on depressive disorder (N women=23,618 and N men=17,336), bipolar disorder (N women=26,573 and N men=22,381), schizophrenia (N women=54,513 and N men=68,287) as SMIs, and oestrogen (N women=229,966 and N men=206,927) and testosterone (N women=230,454 and N men=194,453) as the primary sex hormone traits. Exploratory analyses included other sex hormone traits (progesterone, sex hormone-binding globulin, prolactin, age of menarche, age of menopause). Potential mediators examined included levels of cortisol and CRP, smoking initiation, alcohol dependence, alcohol intake, BMI and body fat distribution. Only individuals of European ancestry were included.

**Results:** We found a widespread pattern of significant genetic correlations between oestrogen/testosterone levels and depressive disorder/schizophrenia, in both positive and negative directions in both sexes (ranging between -0.22 and 0.13). There was no significant genetic correlation for bipolar disorder. In the MR analyses, evidence for causal effects was largely lacking; however, there was consistent evidence for a causal, increasing effect of testosterone levels on schizophrenia risk in men, completely mediated by CRP. Conversely, there was evidence for a causal, increasing effect of liability to schizophrenia on testosterone levels in both sexes.

**Conclusion:** This study offers new insights into the complex aetiology of SMI by comprehensively mapping genetic associations with sex hormone traits, emphasizing the need to further investigate sex hormones’ impact on SMI using larger and more precisely phenotyped samples to identify individuals particularly vulnerable to hormonal disturbances.

## Introduction

Severe mental illness (SMI) forms a leading cause of disease burden and disability.^1^ Despite important developments in psychotherapies and pharmacological treatments of schizophrenia, bipolar disorder, major depressive disorder and their related spectrum disorders, the worldwide burden remains high with little evidence of effective preventive measures.^2^ Prominent sex differences exist in SMI,^3,4^ and increasing evidence points towards a pivotal role for sex hormones.^5^ Elucidation of the exact role of sex hormones in SMI is crucial to tailor sex-specific prevention and treatment.

Oestrogens, the main sex hormone in women, exist as different types with the most important being estrone (E1), oestradiol (E2, or 17b-oestradiol), and oestriol (E3)^6^. Of these, oestradiol is the most potent form. In women, levels of oestrogens vary during the reproductive period, along with the different phases of the menstrual cycle. In the second phase of the menstrual cycle, also referred to as the luteal phase, oestrogens levels decrease. The menopause is characterised by a steep drop in oestrogens. Other hormones that play an important role in reproductive health of women include progesterone, testosterone, and prolactin. In men, testosterone plays a pivotal part in sexual development and behaviours. While testosterone levels vary somewhat over the course of a day, these variations are not equivalent to the hormonal cycle observed in women. Instead, testosterone levels are more influenced by external factors such as lifestyle, as well as by age^7^. In particular, from the age of 40, testosterone levels generally start to decline in men, but the decrease is much more gradual than the hormonal drop during menopause. In both sexes, oestrogens and testosterone are largely bound to Sex-Hormone Binding Globuline (SHBG) in the blood.

There is a clear association between sex hormone levels and the prevalence of SMI. Women show an increased vulnerability for mood disorders and psychosis in periods when oestrogen levels are declining, such as postpartum and perimenopause.^8,9^ For schizophrenia, in addition to a peak of incidence in young adulthood in men, women show more stable incidence rates with a relatively higher perimenopausal onset compared to men of similar ages.^10^ The influence of sex hormones on SMI in men has been less consistently demonstrated, however, testosterone replacement therapy has been shown to sometimes reduce depressive symptoms.^11^

One possible explanation for the associations between SMI and sex hormone levels is an overlapping genetic vulnerability. Twin studies have shown SMIs to be moderately to highly heritable, with estimates ranging between 37% for major depressive disorder and 81% for schizophrenia.^12^ The heritability of sex hormone traits has been less studied; the latest twin study sets testosterone heritability at 12%–51% in women and around 60% in men, and oestradiol heritability at 44% in men and 6%–31% in women.^13^ It is unknown whether these genetic factors overlap, such that the genetic variants that increase risk of SMI may also affect hormone levels. Genome-wide association studies (GWASs) offer a method to explore genetic links between traits, by identifying common genetic variations statistically associated with a specific trait by analysing DNA from many individuals.^14^ These genetic variants can then be correlated between two GWASs resulting in genetic correlations.^15^ Although no studies have found significant genetic correlations between SMIs and sex hormones, recent findings indicate a significant genetic correlation (rg = 0.23) between depression and menopause symptoms.^16^

Another plausible explanation for the link between sex hormones and SMI might be a causal relationship, potentially bidirectional. Possible causal pathways include direct hormonal effects on the brain (e.g. oestrogen modulation of dopamine receptors), mediation/moderation through inflammation,^17^ and HPA-axis activity.^18,19^ Lifestyle factors are also important to consider, as individuals with SMI are more likely to exhibit unhealthy behaviours such as substance use and poor diet, which might in turn alter sex hormone levels.^20,21^ Present evidence predominantly comes from observational studies, which are susceptible to bias from confounding factors and reverse causality. Mendelian randomization (MR) offers a valuable tool to circumvent these biases and explore causal effects, using genetic variants as proxies for risk factors of interest, resembling the framework of instrumental variable analysis.^22^ Because genetic variants are randomly passed on from parents to offspring, bias from confounders can be avoided, provided core assumptions are met.

In this study, we aim to explore mechanisms that might explain the association between sex hormones and SMI using summary-level GWAS data to compute genetic correlations and perform MR. These complementary genetic methods will help assess potential genetic overlap and determine whether associations observed in previous observational research reflect causal effects.

## Methods

Figure 1 provides an overview of our study design (pre-registered here: https://osf.io/qfcz4). First, genetic correlations between sex hormone traits and SMIs were computed to estimate genetic overlap. Second, MR was performed to assess causal effects of sex hormone traits on SMI, and vice versa. Multivariable MR (MVMR) was applied to explore potential mediators.

**Figure 1:**
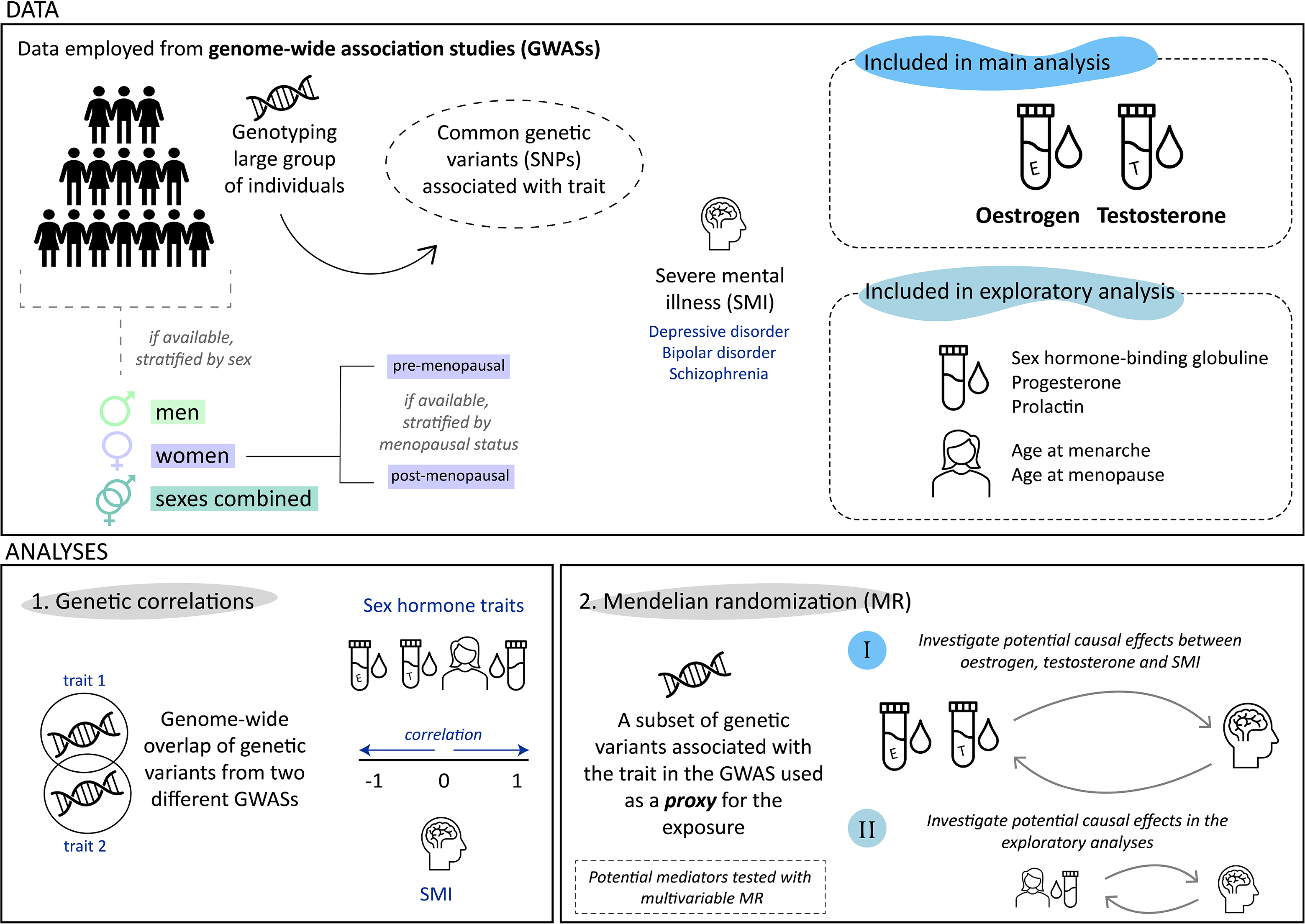
Study overview. This figure outlines the study design (pre-registered here: https://osf.io/qfcz4), which is partitioned into a main analysis focusing on oestrogen and testosterone, and an exploratory analysis including other sex hormones and reproductive traits. Data utilized are derived from GWASs.

### Data

All analyses were performed using publicly available summary statistics from GWAS (see table 1 for a comprehensive overview). For SMI, diagnoses of major depressive disorder, bipolar disorder and schizophrenia-spectrum disorders were included^23-28^. For sex hormone traits, oestrogen and testosterone were included in the main analysis^29,30^, and SHBG, progesterone, prolactin, age of menarche and age of menopause in the exploratory analysis^31-33^. In addition, levels of cortisol and c-reactive protein (CRP), smoking initiation, alcohol dependence, alcohol intake, BMI and body fat distribution were included as potential mediators (see table S1)^34-39^. GWAS were selected with a focus on obtaining large sample sizes to maximize statistical power. In some instances, GWAS with smaller sample sizes, excluding the UK Biobank, were used to avoid sample overlap in the MR analyses, as this could bias the results^40^. For further details on the samples see the supplement.

**Table 1.**
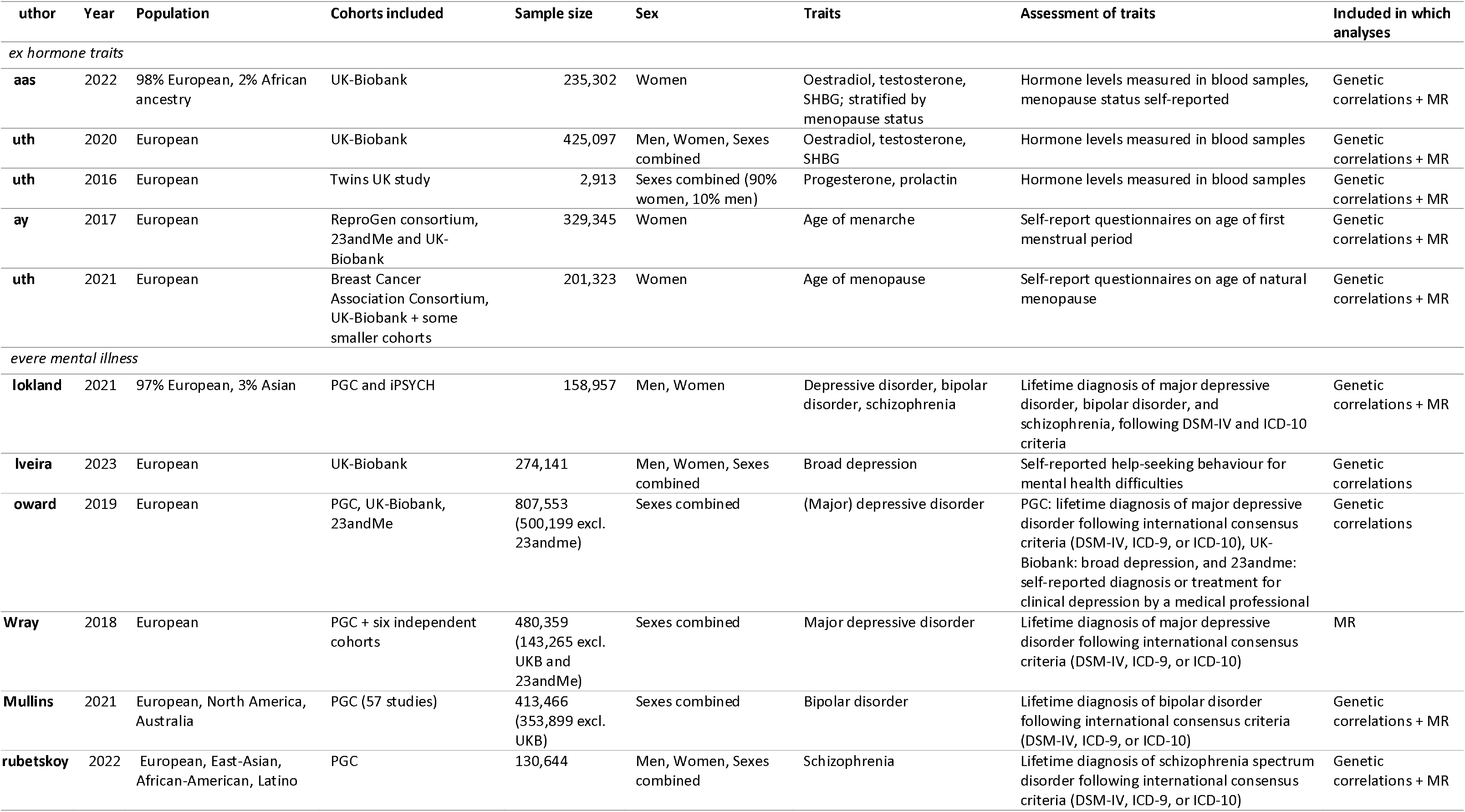
Description of GWAS included.

### Genetic correlations

Genetic correlations were estimated using linkage disequilibrium score (LDSC) regression^15^. LDSC employs summary statistics of genome-wide association studies (GWAS) to estimate the genetic covariation between two traits based on all polygenic effects captured by the genetic variants. In GWAS, the genetic variants generally used are single nucleotide polymorphisms (SNPs), which are variations at a single nucleotide in a DNA sequence. Linkage disequilibrium (LD) represents the degree to which one genetic variant is correlated with a nearby genetic variant within a given population and is used to compute a genetic correlation. The genetic correlation is based on the estimated slope from the regression of the product of z-scores from two GWAS on the LD score. HapMap3 was used as a reference panel for the genome-wide LD scores^41^. We corrected for multiple testing by applying the false discovery rate to the retrieved p-values as described by Benjamini-Hochberg^42^. Estimated genetic correlations can be negative, indicating opposing effects, zero, representing no overlap, or positive, suggesting an overlap in the same direction. An observed correlation could be the result of genuine shared genetic aetiology, meaning that the same genetic variants have a direct influence on different traits. However, an observed correlation could also be the result of confounding factors or causal relationships^43^. The latter was the focus in the subsequent MR analyses, where instead of including SNPs across the whole genome, we selected a small subset of SNPs to proxy an exposure of interest and test causal effects.

### Mendelian randomization

Summary-level MR was applied to investigate potential causal effects. MR uses genetic variants as an instrumental variable, or proxy, for the exposure of interest to investigate whether there are potential causal effects on the outcome of interest. MR relies on three key assumptions that must hold for a MR study to be valid^44^. First, the genetic variants, in this case SNPs, must associate with the exposure of interest (the relevance assumption). Second, the SNPs in the instrument must be independent of confounders (the independence assumption). Third, the SNPs must only affect the outcome through their effect on the exposure of interest (the exclusion restriction assumption). It is important to note that next to these core assumptions, additional assumptions may be relevant, depending on the analysis in question, such as the assumption that the instrumental SNPs for the exposure will predict the exposure in the outcome GWAS sample to the same degree as they do in the exposure GWAS sample (for a complete overview of relevant assumptions see: Richmond et al., 2022^45^). Following MR guidelines^40^, the main effect in the univariable MR analyses was estimated using the Inverse-Variance Weighted (IVW) regression, after which several sensitivity methods were applied with each different assumptions. The IVW-regression is calculated based on the ratio estimates of individual genetic variants, also called Wald ratio, which are the ratios of the variant— outcome association divided by the variant—exposure association. These ratio estimates are then meta-analysed with weights based on their inverse variance. Independent instrument SNPs for the MR analyses were selected based on their genome-wide significant association with the exposure, adhering to a p-value threshold of 5e-08.

We applied 6 sensitivity methods: weighted median, weighted mode, MR-Egger, MR pleiotropy residual sum and outlier (MR-PRESSO), Steiger filtering, and GSMR. Further description of these methods can be found in the supplement. We also performed leave-one-out analyses, repeating IVW after removing each SNP, and computed Cochran’s Q to assess heterogeneity between SNP-estimates in each instrument. To assess instrument strength, we computed the F-statistic (F > 10 is sufficiently strong). Analyses were performed in R (4.3.2), using packages: “TwoSampleMR,” “GSMR,” “psych,” and “MR-PRESSO.”

MR results were considered as robust when findings were consistent across methods. The majority of the sensitivity methods rely on stricter assumptions than IVW, resulting in less power. Therefore, their statistical evidence will be weaker, even for a true causal effect. We consider a finding to be consistent across sensitivity methods if the beta coefficient is in the same direction and of comparable size (or larger). Findings were described as showing no clear evidence, weak evidence, evidence or strong evidence for a causal effect, taking into account IVW and the sensitivity methods, adhering to the broad interpretation of P-values described by Sterne and Davey Smith (2001)^46^. We did not correct for multiple testing in the MR analyses, as this may be excessively cautious, especially considering our focus on relationships with prior epidemiological support. As suggested in the latest MR guidelines, we separated the primary analysis (including solely oestrogen and testosterone), used to answer our hypothesis, from the exploratory analysis^40^. The exploratory analysis investigates other sex hormones and reproductive traits, with less established prior knowledge in relation to SMI.

Using multivariable MR (MVMR), a range of potential mediators were investigated. For each univariable analysis, we added each potential mediator separately to prevent violation of the linearity and homogeneity assumptions^47^. If the direct effect substantially diminished after the addition of the second exposure compared to the effect observed in the univariable MR analysis, we considered this as a mediator. The Sanderson-Windmeijer conditional F-statistic was computed to test instrument strength^48^. To evaluate robustness of the findings multivariable MR-Egger and an adaption of the Cochran’s Q statistic were performed. Again all analyses were performed in R, additionally using the package: “MVMR”.

## Results

### Genetic correlations

Figure 2 presents an overview of genetic correlations between sex hormones and severe mental illnesses. Genetic correlations involving progesterone or prolactin were omitted because the small heritability in the GWASs did not permit us to conduct LD score regression. Out of a total of 45 estimated correlations, 16 were significant, of which 11 remained significant after FDR correction.

**Figure 2:**
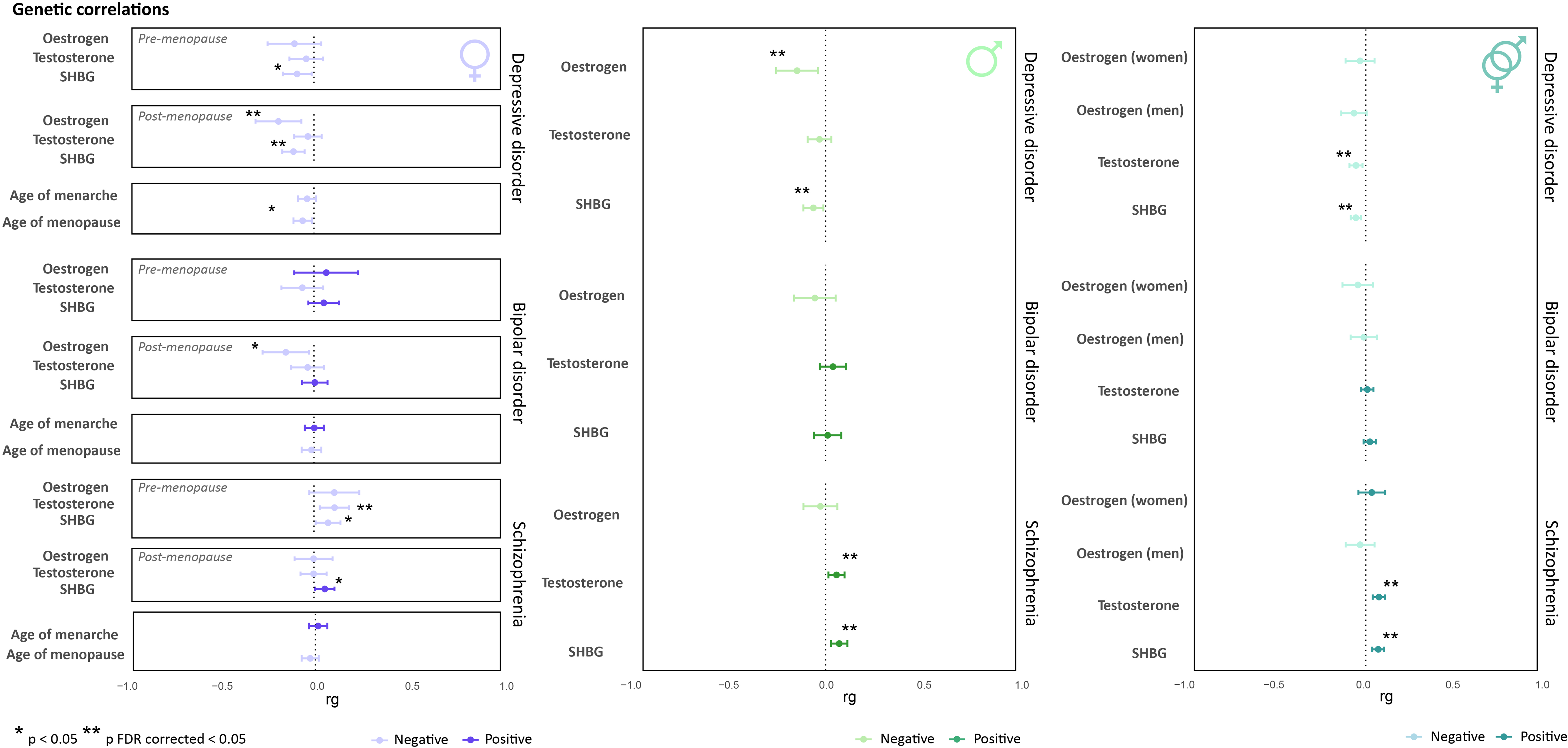
Genetic correlations between sex hormones traits and SMIs. This figure illustrates the genetic correlations between levels of key sex hormones (oestrogen, testosterone) and related sex hormones and reproductive traits (SHBG, and the age of onset for menarche and menopause) in relation to the SMI (from top to bottom: depressive disorder, bipolar disorder, and schizophrenia). Genetic correlations involving progesterone or prolactin were omitted because the small heritability in the GWASs did not permit us to conduct LD score regression. Asterisks denote statistical significance, with * p < 0.05 uncorrected, and ** p < 0.05 FDR corrected.

#### Women

Genetic correlations ranged between -0.22 and 0.13 in women. In depressive disorder, a negative correlation was found with post-menopausal oestrogen levels (rg=-0.22, standard error (SE)=0.07, p_FDR_=0.020), while pre-menopausal oestrogen levels did not show a significant correlation. In addition, SHBG levels demonstrated negative correlations with depressive disorder. In bipolar disorder, there were no significant correlations after FDR adjustment. In schizophrenia, positive correlations were found with pre-menopausal and overall testosterone (pre-menopause rg=0.1298, SE=0.0475, p_FDR_=0.03465; overall rg=0.0779, SE=0.024, p_FDR_=0.0132).

#### Men

Genetic correlations ranged between -0.16 and 0.08 in men. In depressive disorder, a negative correlation was found with oestrogen levels (rg=-0.16, SE=0.06, p_FDR_=0.025) and SHBG levels (rg=-0.07, SE=0.03, p_FDR_=0.037). In bipolar disorder, there were no significant correlations after FDR adjustment. In schizophrenia, there was a positive correlation with testosterone (rg=0.06, SE=0.02, p_FDR_=0.0249) and SHBG levels (rg=0.08, SE=0.02, p_FDR_=0.012).

#### Sexes combined

Genetic correlations ranged between -0.07 and 0.08 in sexes combined. In depressive disorder, a negative correlation was observed with testosterone (rg=-0.06, SE=0.02, p_FDR_=0.010) and SHBG levels (rg=-0.06, SE=0.02, p_FDR_=4E-04). Again, there were no significant correlations after FDR adjustment in bipolar disorder. In schizophrenia, a positive correlation was found with testosterone (rg=0.08, SE=0.02, p_FDR_=2.4E-04) and SHBG levels (rg=0.08, SE=0.02, p_FDR_=2.4E-04).

### Mendelian randomization analyses

All instruments employed in the univariable MR analyses were sufficiently strong (see table 2 for an overview). Main results are depicted in figure 3 and table S3. GSMR results, which were consistently almost equivalent to the IVW results, are reported in table S5.

**Table 2.** Description of MR instruments.

| Phenotype | Stratified on menopausal status | GWAS | Measures | Sample size | SNPs, N | F statistic | I2GX* |
| --- | --- | --- | --- | --- | --- | --- | --- |
| **Instruments used for women only** |  |  |  |  |  |  |  |
| *Sex hormone traits* |  |  |  |  |  |  |  |
| **Oestrogen** | Pre-menopausal | Haas, 2022 | Detectable concentrations yes/no | 51,081; 33,032 cases, 18,049 controls | 4 | 50 – 54 | 0.94 – 0.95 |
|  | Post-menopausal | Haas, 2022 | Detectable concentrations yes/no | 84,194; 3,759 cases, 80,435 controls | 6 | 55 – 67 | 0.95 – 0.98 |
|  | Not stratified | Haas, 2022 | Detectable concentrations yes/no | 229,966; 48,487 cases, 181,479 controls | 10 | 51 – 68 | 0.90 – 0.95 |
| **Testosterone** | Post-menopausal | Haas, 2022 | ng/dL | 84,003 | 2 | 32 | N.A. |
|  | Not stratified | Haas, 2022 | ng/dL | 230,454 | 254 | 93 – 107 | 0.96 – 0.97 |
| **SHBG** | Pre-menopausal | Haas, 2022 | ng/dL | 43,477 | 31 | 71 – 73 | 0.91 – 0.94 |
|  | Post-menopausal | Haas, 2022 | ng/dL | 92,911 | 103 | 67 – 70 | 0.90 – 0.93 |
| **Progesterone** | Not stratified | Ruth, 2016 | nmol/L | 2,689 | 2 | 52 | 0.37 |
| **Age at menarche** | Not stratified | Day, 2017 | years | 329,345 | 389 | 65 – 69 | 0.89 – 0.92 |
| **Age at menopause** | Not stratified | Ruth, 2021 | years | 201,323 | 290 | 99 – 103 | 0.95 – 0.96 |
| *SMI* |  |  |  |  |  |  |  |
| **Schizophrenia** | Not stratified | Trubetskoy, 2022 | Lifetime diagnosis yes/no | 54,513; 17,710 cases, 36,803 controls | 22 | 37 | 0.31 – 0.91 |
| **Instruments used for men only** |  |  |  |  |  |  |  |
| *Sex hormone traits* |  |  |  |  |  |  |  |
| **Oestrogen** | Not stratified | Ruth, 2020 | Detectable concentrations yes/no | 206,927; cases and controls unknown | 22 | 79 – 87 | 0.94 – 0.95 |
| **Testosterone** | Not stratified | Ruth, 2020 | nmol/L | 194,453 | 233 | 76 – 114 | 0.93 – 0.97 |
| **SHBG** | Not stratified | Ruth, 2020 | nmol/L | 180,726 | 276 | 102 – 110 | 0.95 – 0.96 |
| *SMI* |  |  |  |  |  |  |  |
| **Schizophrenia** | Not stratified | Trubetskoy, 2022 | Lifetime diagnosis yes/no | 68,287; 33,097 cases, 35,190 controls | 56 | 42 | 0.57 |
| **Instruments used for sexes combined** |  |  |  |  |  |  |  |
| *Sex hormone traits* |  |  |  |  |  |  |  |
| **Testosterone** | Not stratified | Ruth, 2020 | nmol/L | 425,097 | 238 | 132 – 133 | 0.97 |
| **SHBG** | Not stratified | Ruth, 2020 | nmol/L | 370,125 | 466 | 107 – 109 | 0.96 |
| *SMI* |  |  |  |  |  |  |  |
| **Depressive disorder** | Not stratified | Wray, 2018 | Lifetime diagnosis yes/no | 143,265; 45,591 cases, 97,674 controls | 2 | 36 | 0 |
| **Bipolar disorder** | Not stratified | Mullins, 2021 | Lifetime diagnosis yes/no | 353,899; 40,463 cases, 313,436 controls | 51 | 39 | 0.35 |
| **Schizophrenia** | Not stratified | Trubetskoy, 2022 | Lifetime diagnosis yes/no | 130,644; 53,386 cases, 77,258 controls | 185 | 44 | 0.60 |

\*The I2GX statistic was computed to assess whether the NOME assumption was satisfied for MR-Egger. A I2GX statistic higher than 0.9 indicates that bias due to violation of the NOME assumption is not likely. A I2GX statistic between 0.6 and 0.9 suggests bias can be corrected for with simulation extrapolation (SIMEX) MR-Egger.

**Figure 3:**
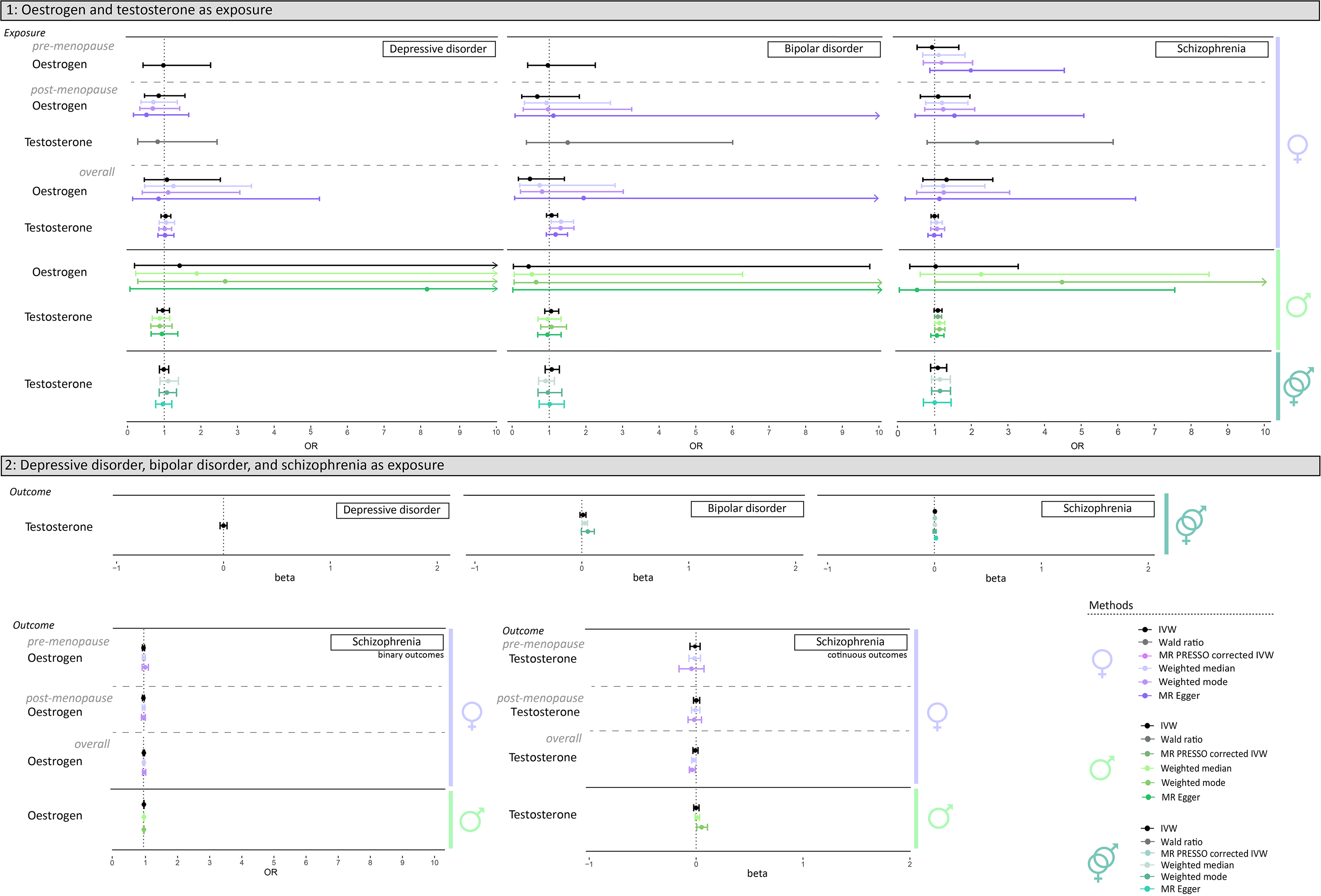
Main Mendelian randomization analyses. Panel 1 shows plots of the main MR results including oestrogen and testosterone levels as exposures, while panel 2 shows plots of the main MR results in the reverse direction, with depressive disorder, bipolar disorder, and schizophrenia as exposures. Analyses for women are depicted in purple, men in green, and combined sexes in blue. Women’s analyses are further stratified by menopausal status: pre-menopause, post-menopause, and overall. Separate graphs are used for binary and continuous outcomes due to the difference in outcome measures. On the right, the legend displays the methods used: Inverse-Variance Weighted (IVW), Wald ratio, MR PRESSO corrected IVW, weighted median, weighted mode, and MR-Egger. OR = odds ratio.

#### Women

There was no evidence supporting causal effects of oestrogen or testosterone levels on SMI in women. In the reverse direction, there was no evidence supporting causal effects of liability to mental illness on oestrogen or testosterone levels in women.

#### Men

There was weak evidence for an increasing effect of higher testosterone levels on schizophrenia risk (OR_IVW_=1.09, 95%CIs=0.98 to 1.2, p=0.099). Weighted median and weighted mode confirmed this effect with stronger statistical evidence. The MR-Egger intercept did not show evidence for horizontal pleiotropy (p=0.721), and its slope corroborated the direction, with a small decrease in effect size (OR=1.06, 95% CIs=0.9 1.26, p=0.486). There was strong evidence for heterogeneity (Qp=8E-17). MR-PRESSO detected 5 outliers, and removal of these outliers increased the statistical evidence (p=0.057). Leave-one-out analysis also demonstrated that rs113017476 and rs12406721 suppressed the effect. The SNP rs113017476 was shown to have a negative effect on schizophrenia (IVW Wald ratio: OR=0.60, 95%CIs=0.88 to 0.41, p=0.010) and is mapped to the SRD5A2 gene, a gene involved in processing androgens. Steiger filtering removed two other SNPs, similarly resulting in stronger evidence (table S5). In the MVMR analyses, the adjustment for CRP decreased the effect with ∼109% (IVW adjusted for CRP: OR=0.99, 95%CIs=0.83 to 1.18, p=0.932; table S8), with evidence for a negative effect of CRP on schizophrenia (OR_IVW_=0.88, 95%CIs=0.80 to 0.96, p=0.003). The Q-statistic indicated heterogeneity (p= 4.7E-10, table S12), but the MR-Egger intercept showed no evidence for pleiotropy (table S14). Conversely, there was no evidence for causal effects of liability to SMI on oestrogen or testosterone levels in men.

#### Sexes combined

There was no evidence supporting causal effects of oestrogen or testosterone levels on SMI risk in both sexes. In the reverse direction, there was weak evidence for an increasing effect of liability to schizophrenia on testosterone levels (beta_IVW_=0.01, 95%CIs= -5.4E-04 to 0.01, p=0.076). Weighted mode and weighted median regression confirmed the direction of effect with less statistical evidence. The MR-Egger intercept did not show evidence for horizontal pleiotropy (p=0.561). However, the MR-Egger regression slope did not confirm the direction (beta=-1.5E-03, 95%CIs=-0.02 to 0.02, p=0.901), although this could potentially be due to limited power. There was strong evidence for heterogeneity (Qp=6.7E-38). MR-PRESSO removed 8 outliers, which increased the statistical evidence (p=0.013). Leave-one-out analysis showed that rs11191580 and rs79780963 (both mapped to the NT5C2 gene) supressed the effect. Steiger filtering did not remove any SNPs. Adjusting for potential mediators with MVMR did not have a significant impact; however, the instruments for cortisol and alcohol intake lacked sufficient instrument strength (table S11), potentially limiting our ability to detect an effect.

#### Exploratory analyses

Results of the exploratory analyses are presented in the supplement. In brief, consistent evidence was found in women for an increasing effect of pre-menopausal and post-menopausal SHBG levels on depressive disorder risk, and conversely, for an increasing effect of liability to schizophrenia on post-menopausal SHBG levels. In men, there was no evidence for causal effects. In sexes combined, there was consistent evidence for an increasing effect of liability to schizophrenia on SHBG levels, and for an increasing effect of liability to bipolar disorder on SHBG levels. In the MVMR analyses, CRP was shown to completely mediate the effect of liability to bipolar disorder on SHBG levels (∼109% decrease).

## Discussion

The aim of this study was to investigate the association between levels of sex hormones and SMI risk using genetic-informed approaches. To assess whether there might be a shared genetic aetiology, we computed genetic correlations of oestrogen, testosterone, and related sex hormone traits with depressive disorder, bipolar disorder, and schizophrenia. The results showed a widespread pattern of significant correlations between oestrogen and testosterone levels and depressive disorder and schizophrenia, in both positive and negative directions in both sexes (ranging between -0.22 and 0.13). In addition, we performed MR analyses to explore causality. There was consistent evidence for a causal, increasing effect of testosterone levels on schizophrenia in men, completely mediated by CRP. Conversely, there was evidence for a causal, increasing effect of schizophrenia liability on testosterone levels in both sexes.

The substantial genetic correlations found between sex hormones and SMI suggest an overlap in genetic aetiology; a novel finding. Both positive and negative correlations were observed, reflecting the complex relation between sex hormones and SMI, as seen in non-genetic studies.^49^ Notably, depressive disorder showed only negative correlations with sex hormones, while schizophrenia showed only positive correlations, indicating distinct hormonal patterns. It should be noted that genetic correlations are merely a correlation between two GWASs, and apart from shared genetic aetiology, there are multiple mechanisms that could underly this observed correlation.^43^ The only study that computed similar genetic correlations, including testosterone and SMI, showed results close to zero, most likely due to smaller sample sizes.^50^ Given the current abundance of GWAS data, the lack of studies exploring genetic correlations between sex hormones and SMI highlights the significant gap in research investigating these relations, especially considering the well-established observational clinical correlation. Future research should further investigate these genome-wide genetic correlations at a local level, potentially revealing diverse correlations within different genomic regions, thereby improving our understanding of the intricate dynamics of hormonal functioning in SMI.

A surprising finding in the MR analyses was the increasing effect of testosterone levels on schizophrenia in men. The research field investigating testosterone in schizophrenia is still in a preliminary stage, characterized by a scarcity of studies and the necessity for more robust methodologies and insights. Acknowledging the contradictive outcomes and typically heterogeneous samples of existing studies, the majority of evidence nonetheless indicates either lower levels of testosterone or no significant difference compared to healthy controls,^51^ which contrasts with our findings. However, a meta-analysis revealed elevated testosterone levels in male schizophrenia patients experiencing acute relapse, and in patients experiencing their first episode,^52^ suggesting a potential interplay with stage of illness. Furthermore, Schwarz et al.^53^ identified a subgroup of schizophrenia patients with hormonal abnormalities, including increased testosterone. It has been hypothesized that testosterone might be protective because of its anti-inflammatory properties,^54^ aligning with the growing recognition of inflammation in mental illness.^55^ Our MVMR analysis confirmed the anti-inflammatory effect of testosterone, as CRP’s effect decreased. However, CRP was found to be protective against schizophrenia risk, a finding corroborated in the GWAS employed for CRP^35^ and another MR study.^56^ The authors hypothesize that this may be due to a weaker immune response at birth that has been observed in schizophrenia patients. It is important to consider that in the current study, MR analyses investigate the effect of a *life-long* exposure, which disregards the time-dependent effects. Additionally, the interlinked nature of testosterone’s biological functions, influenced by its conversion to oestrogens and interactions with various hormonal systems,^57^ makes it difficult to be fully represented by the genetic variants employed in the current study. Furthermore, heterogeneity was shown to be present and leave-one-out analysis uncovered a SNP involved in processing androgens to have a strong negative effect. Therefore, these results warrant cautious interpretation.

We did not find evidence for a causal link between lower oestrogen levels and SMI risk, contrary to the widespread belief that oestrogen plays a neuroprotective role.^58^ Prior studies have consistently shown lower oestrogen levels in women with SMI, recently supported by randomized-controlled trials that show administering exogenous oestrogen is effective in treating SMI.^59,60^ A possible explanation for our results could be that our MR analysis lacked sufficient power. The instruments for oestrogen contained only two to four SNPs stratified on menopausal status, up to a maximum of eight not stratified, and showed heterogeneity. Given the complex and multifactorial nature of SMI, any true causal effect of oestrogen, if present, is likely to be modest and therefore challenging to detect. It is plausible that hormonal imbalances may only affect a specific, vulnerable subgroup within the broader SMI population. Additionally, we had to employ a binary measure of oestrogen (above/below detection limit) that does not fully capture the variation in oestrogen levels, potentially limiting our ability to detect true effects. Nevertheless, our study did uncover negative genetic correlations of post-menopausal oestrogen levels with depressive and bipolar disorder. Interestingly, these correlations were more pronounced depending on menopausal status, aligning previous research showing associations between lower levels of oestrogen after menopause and increased depressive disorder risk.^61^ We also observed consistent evidence in women for causal effects of SHBG on depressive disorder, and conversely, of schizophrenia on SHBG, which carries oestrogen. While some studies corroborate the positive association of SHBG levels with SMI^62-64^, interpreting these findings is complex due to SHBG’s interaction with various hormones and other factors, such as energy balance and physical activity.^65^

The strengths of this study include the extensive sex-specific investigations of the main sex hormones and related traits, stratified by menopausal status, in relation to SMI. To our knowledge, this is the first study to perform such a thorough genetic investigation. There were also some limitations. First, the genetic variants for sex hormones were based on single blood measurements, which might not fully capture hormone variability over time, especially in women due to menstrual cycles, pregnancy, and menopause. This might affect the accuracy of genetic associations across different life stages or physiological conditions. Second, some analyses lacked power due to small GWAS sample sizes for prolactin, progesterone, and cortisol. Additionally, to avoid sample overlap, certain analyses could not be performed or consisted of smaller sample sizes. Third, it is likely that relatively well-functioning mental illness patients were included in the GWAS, leading to potential selection bias. Fourth, potential biases such as dynastic effects, assortative mating, and population stratification could influence both genetic correlations and MR.^66^

This study offers new insights into the complex aetiology of depressive disorder and schizophrenia by comprehensively mapping genetic associations with sex hormone traits. Our findings, highlighting CRP as a mediator, emphasize the importance of further investigating sex hormones within the multifactorial model of mental illness, including inflammation. Future research should build upon these findings using larger, precisely phenotyped samples, measuring sex hormones at multiple time-points in the cycle, and focus on the identification of individuals particularly vulnerable to hormonal disturbances. There is an underrepresentation of genetic data on female sex hormones which needs to be addressed in future samples. Together, these steps will facilitate more precise and mechanistic findings and the development of tailored treatment strategies.

## Data Availability

All analyses in this study employed publicly available summary-level GWAS data

