## Supplement for "The role of sex hormones in severe mental illness: a genetic exploration"

### Supplementary methods

#### Biological sex versus gender

In the current study we focus specifically on biological sex, not gender. This distinction is made because we are examining the relation between sex hormones and SMI. Biological sex, defined by the presence of X and Y chromosomes, directly influences sex hormone production, which is central to our analyses. We acknowledge that this focus on biological sex does not fully encompass the complexities of gender identity, which is equally significant but beyond the scope of our hormonal study.

#### GWAS data on SMI

To study the relation between sex hormones and severe mental illness (SMI), we included the diagnoses major depressive disorder, bipolar disorder, and schizophrenia. It should be noted that the selection of a genome-wide association study (GWAS) varied depending on the analysis type. For genetic correlations, we utilized the largest available GWAS, including UK-Biobank data. For the mendelian randomization (MR) analyses we excluded UK-Biobank or selected an alternative GWAS without UK-Biobank, to avoid sample overlap^1^, as our instruments for sex hormone traits consisted exclusively of UK-Biobank. Additionally, separate GWASs were sometimes necessary for the sex-stratified analyses and for the analyses with sexes combined.

Sexes combined

For depressive disorder, we selected the largest GWAS available on depression including UK-Biobank for the genetic correlations, a meta-analysis of Howard et al.^2^ including the Psychiatric Genomics Consortium (PGC) and 23andMe cohorts with a total of 807,553 individuals (246,363 cases and 561,190 controls). 23andme was excluded, resulting in a final sample size of 500,199 (170,756 cases and 329,443 controls). The PGC cohorts defined depressive disorder as a lifetime diagnosis of major depressive disorder following international consensus criteria (DSM-IV, ICD-9, or ICD-10). In UK-Biobank, the broad definition for depression was used, defined as a yes answer to one of the following questions: ‘Have you ever seen a general practitioner for nerves, anxiety, tension or depression?’ or ‘Have you ever seen a psychiatrist for nerves, anxiety, tension or depression?’. 23andMe used a self-reported measure indicating a diagnosis or treatment for clinical depression my a medical professional. The heritability was estimated at 8.9% (standard error (SE)=0.003). For the MR analyses on depressive disorder, the PGC GWAS of Wray et al.^3^ was selected, a meta-analysis including 480,359 participants (135,458 cases and 344,901 controls). UK-Biobank and 23andme were excluded, resulting in a sample size of 143,265 individuals in the current study (45,591 cases and 97,674 controls). For bipolar disorder, we utilized the GWAS of the PGC Bipolar Disorder Working Group^4^ for analyses in sexes combined. The GWAS included a meta-analysis combining 57 studies from 21 countries in Europe, North America and Australia, with a total of 413,466 participants (41,917 cases and 371,549 controls). Summary-statistics stratified on sex were not available. For the MR analyses, UK-Biobank was excluded, resulting in a sample size of 353,899 (40,463 cases and 313,436 controls). Cases were defined as a lifetime diagnosis of bipolar disorder, following international consensus criteria (DSM-IV, ICD-9, or ICD-10) and assessed with the use of structured diagnostic instruments, checklists administered to clinicians, or reviews of medical records. Heritability was estimated at 18.6% (SE=0.008). For schizophrenia, the PGC GWAS of 2022^5^ was employed, including 175,799 participants (74,776 cases and 101,023 controls). Only individuals of European ancestry were included in this study, resulting in a final sample size of 130,644 participants (53,386 cases and 77,258 controls). Cases were defined as individuals diagnosed with a schizophrenia spectrum disorder based on the DSM-IV criteria. Sex-stratified data were also available. The heritability was estimated to be 24% (SE=0.007).

Sex-stratified

The sex-stratified GWAS for SMI performed in the study of Blokland et al.^6^ was included. The sex-stratified GWAS for both depressive disorder (N = 40,954; 15,970 cases and 24,984 controls) and bipolar disorder (N = 48,954; 18,958 cases and 29,996 controls) were used in the current study. The sample of depressive disorder consisted of 26,573 women and 22,381 men, and the sample of bipolar disorder of 23,618 women and 17,336 men. Cases were defined as a lifetime diagnosis of the disorder, based on clinical DSM-IV or ICD-10 interviews. The East-Asian cohorts were excluded. For the genetic correlations, the largest available sex-stratified GWAS with UK-Biobank on depressive disorder was also included (N = 274,141; 177,152 cases and 96,989 controls), performed by Silveira and colleagues^7^. The sample compromised 146,274 women and 127,867 men, solely of European ancestry. The broad definition of depression in UK-Biobank was used, described as “self-reported past help-seeking for problems with nerves, anxiety, tension or depression”^2^. For schizophrenia, the GWAS of the PGC^5^ was employed, the same as in the analyses with sexes combined. Again, only individuals of European ancestry were included, resulting in a total sample size of 54,513 for women and 68,287 for men.

#### GWAS data on sex hormone traits

The GWAS of Haas and colleagues^8^ was used for the sex hormones oestrogen, testosterone, and SHBG, in women only. UK-Biobank data of blood samples were used measuring circulating concentrations of oestradiol (field 30800-0.0), testosterone (field 30850-0.0) and SHBG (field 20830-0.0), stratified on menopausal status. Pregnant women were excluded. In the post-menopausal sample, women reporting any use of hormone replacement therapy were also excluded. Individuals of non-European ancestry were excluded for the current study, resulting in a final sample size of 229,966 women (62,587 pre-menopausal and 124,820 post-menopausal). The heritability of oestrogen was estimated as 1.5% (SEM=0.2) for the binary trait and 1.1% (SEM = 1.0) for the continuous measure. When stratified on menopausal status, the heritability of binary oestrogen increased to 3.9 (SEM=1.0) for pre-menopausal and 3.5 (SEM=0.6) for post-menopausal. The heritability of testosterone was estimated as 11% (SEM=0.9) and stratifying did not significantly change this. The heritability of SHBG was estimated as 19% (SEM=1.9), and when stratified as 23% (SEM=2.8) for pre-menopausal and 22% (SEM=2.6) for post-menopausal. The GWAS of Ruth and colleagues from 2020^9^ was employed for oestrogen, testosterone, and SHBG in men. Data was also available in sexes combined, except for oestrogen. The total sample consisted of 425,097 individuals of European ancestry. Again, UK-Biobank data was employed, including concentrations of oestrogen, testosterone, and SHBG measured in blood samples. Data was not available stratified on menopausal status. The heritability was estimated as 17% (standard error of the mean (SEM) = 1,2) in men and 13% (SEM = 0.8) in women for testosterone, 2% (SEM = 0.4) in men and 1.6 (SEM = 1) in women for oestradiol, and 21% (SEM = 1.2) in men and 20% (SEM = 0.1) in women for SHBG. For progesterone and prolactin, the GWAS of Ruth and colleagues from 2016^10^ was employed, including 2,914 individuals of European ancestry (up to 294 men). Circulating concentrations of sex hormones were measured in blood samples. Individuals who were pregnant, who were taking oral contraceptives, and who were currently receiving hormone replacement therapy were excluded. The sex hormones were standardised for sex, age, BMI, menopausal status, and state of menstrual cycle. The effect sizes were estimated as a per-allele standard deviation change in the covariate-adjusted transformed residuals. For age of menarche, the GWAS of Day and colleagues from 2017^11^ was used, including 329,345 women of European ancestry. Age at menarche was self-reported, and women who reported an age of menarche lower than 9 years or higher than 17 years were excluded. Birth year was included as a covariate. Heritability was reported around 25%; 28.8% (SE=2.3%) for early age at menarche and 21.5% (SE=2.5%) for late age at menarche. For age of menopause, the GWAS of Ruth and colleagues from 2021^12^ was utilized, including 201,323 women of European ancestry. Age at natural menopause was self-reported and defined as the age at last menstrual period, succeeded by 12 months of amenorrhea. Women with a natural menopause from age of 40 to 60 were included. Women with a menopause caused by hysterectomy, bilateral ovariectomy, radiation or chemotherapy were excluded, and women who used hormone replacement therapy before menopause.

#### GWAS on mediators

For the potential mediators included in the multivariable MR (MVMR) analyses, we selected the largest available GWASs excluding UK-Biobank for cortisol^13^, CRP^14^, alcohol intake^15^, alcohol dependence^16^, smoking initiation^15^, BMI^17^, and body fat distribution^18^. See table S1 for more information on these GWASs.

| Author | Year | Population | Cohorts included | Sample size | Sex | Traits | Assessment of traits |
| --- | --- | --- | --- | --- | --- | --- | --- |
| Crawford | 2021 | European | CORtisol NETwork (CORNET) consortium | 25,314 | Sexes combined | Cortisol | Measured morning plasma cortisol |
| Ligthart | 2018 | European | CHARGE consortium (HapMap) | 204,402 | Sexes combined | CRP | Serum CRP in mg/L using standard laboratory techniques |
| Saunders | 2022 | Multi-ancestry  (2,669,029 European ancestry) | GSCAN | 3,383,199 | Sexes combined | Alcohol intake (drinks per week), smoking initiation | Alcohol intake was defined as the average number of drinks a participant reported drinking each week, and smoking initiation was defined as ever being a regular smoker in their life (current or former) |
| Walters | 2018 | Mutli-ancestry (46,568 European ancestry) | PGC (28 cohorts) | 52,848 | Sexes combined | Alcohol dependence | DSM-IV diagnosis for alcohol dependence (or, for one cohort, DSM-III), by clinician ratings or semi-structured interviews |
| Locke | 2015 | Multi-ancestry (322,154 European ancestry) | 125 studies | 339,224 | Sexes combined | BMI | Measured or self-reported weight in kg per height in metres squared |
| Shungin | 2015 | European | GIANT consortium | 210,088 | Men, Women, Sexes combined | Body fat distribution (Waist-Hip ratio) | The ratio of waist and hip circumference, without adjustment for BMI |

**Table S1.** Overview of GWASs used for the potential mediators included in the MVMR analyses.

#### Instrument selection

We selected SNPs as instrumental variables if they met the genome-wide significance threshold (p < 5e-08). We ensured that only independent SNPs were used, which was assessed based on the identification in the GWAS itself. If the independent instrument SNPs were not available in the GWAS, we performed linkage disequilibrium (LD) clumping with a threshold of r2 = 0.01 to establish SNP independence. Following the MR guidelines, we selected the unadjusted phenotypes, to minimize risk of collider bias^19^.

Our analyses were performed both sex-stratified and with sexes combined. Additionally, for women, we further stratified the analyses based on menopausal status when data on this was available. In some instances, instruments stratified by sex contained insufficient or no SNPs. For the mental illness phenotypes, we prioritized adherence to the strict genome-wide significance, due to the less understood biological bases of these phenotypes. As a result, some relations could not be tested in the reverse direction. For a detailed understanding of our instrument selection process, see the decision trees provided in figures S1 and S2.

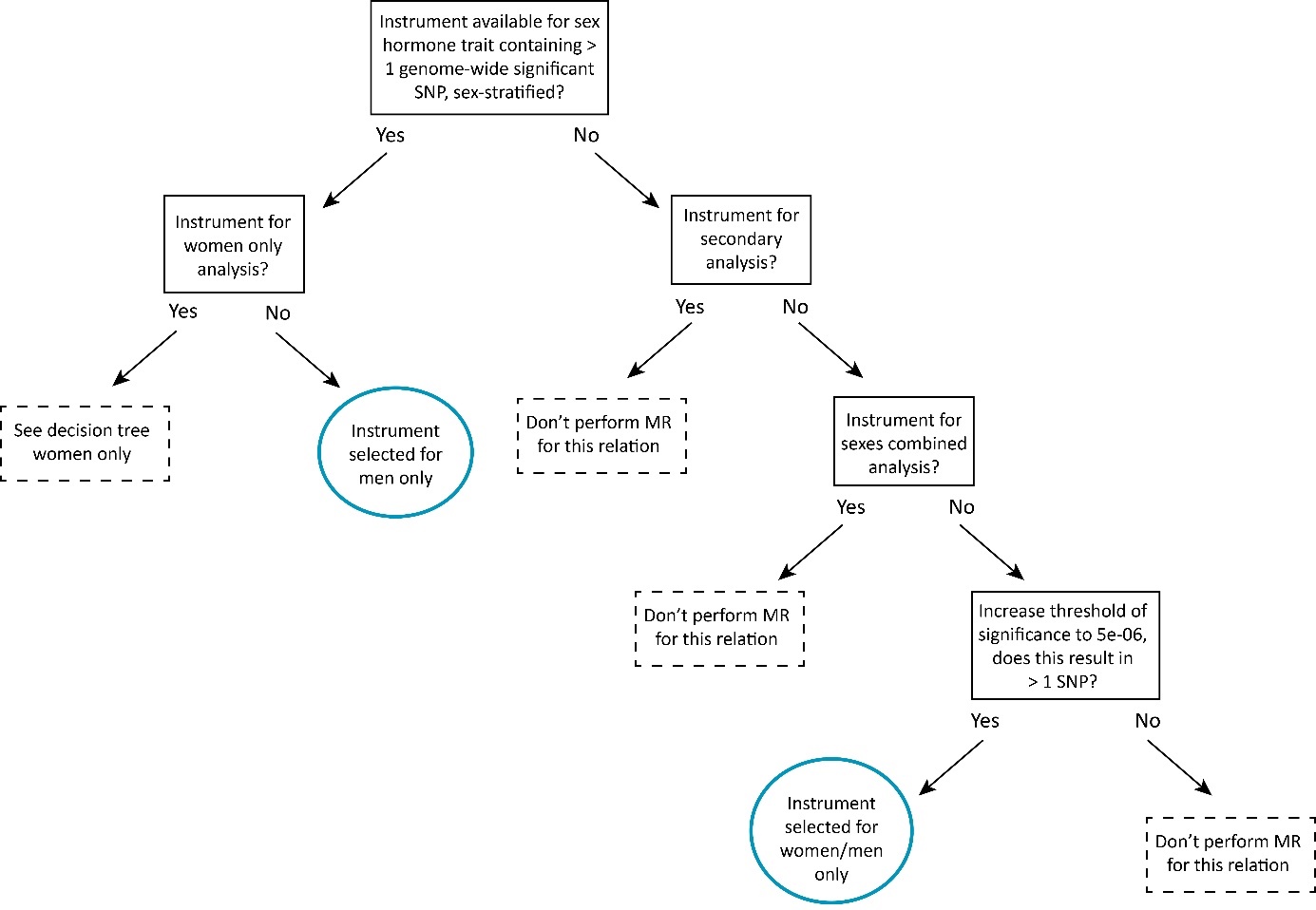

**Figure S1.** Decision tree for selecting instruments for the sex hormone traits

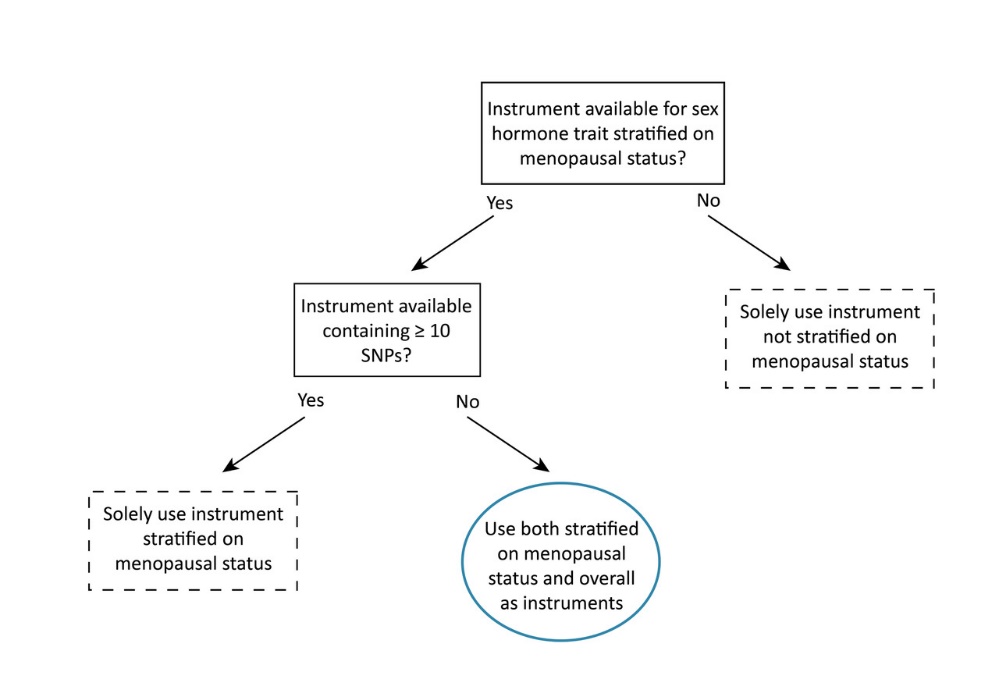

**Figure S2.** Decision tree for selecting instruments for the sex hormone traits in women only

#### Sensitivity analyses

First, we applied the basic MR sensitivity approaches, including weighted median^20^, weighted mode^21^, and MR-Egger^22^. Weighted median and weighted mode regression offer alternative estimates compared to the IVW regression, ensuring a reliable outcome even if some of the instrumental SNPs are invalid. MR-Egger explicitly tests for horizontal pleiotropy, by estimating the intercept is freely which provides an estimation of the average horizontally pleiotropic effect. The MR-Egger intercept then indicates evidence for potential pleiotropy, using a p-value cut-off of 0.05. MR-Egger depends on two assumptions: the INstrument Strength Independent of Direct Effect (InSIDE) assumption and the NO Measurement Error (NOME) assumption. The InSIDE assumption states that any pleiotropic effects present are not correlated with the instrument strength. It should be noted that MR-Egger has markedly lower statistical power compared to the IVW.

If the IVW indicated evidence of a causal effect, we applied three additional sensitivity analyses: MR pleiotropy residual sum and outlier (MR-PRESSO) analysis^23^, Steiger filtering^24^, and generalized summary data-based MR (GSMR)^25^. MR-PRESSO tests and corrects for horizontal pleiotropic outliers. Steiger filtering was applied corrects for reverse causality, through the identification of SNPs that explain a larger amount of variance in the outcome compared to the exposure, and excluding those. GSMR is a method with higher statistical power, achieved by using low levels of LD between the included SNPs. GSMR also performs a filtering step (HEIDI filtering) in which it removes outliers. Analyses were performed in R (4.3.2), using packages: “TwoSampleMR,” “GSMR,” “psych,” and “MR-PRESSO”.

Multivariable MR

For each univariable analysis, we added each potential mediator separately to prevent violation of the linearity and homogeneity assumptions^26^. If the direct effect substantially diminished after the addition of the second exposure compared to the effect observed in the univariable MR analysis, we considered this as a mediator. The Sanderson-Windmeijer conditional F-statistic was computed to test instrument strength^27^. To evaluate robustness of the findings multivariable MR-Egger and an adaption of the Cochran’s Q statistic were performed. Again, analyses were performed in R, using the package “MVMR”.

#### STROBE-MR Guidelines

Official guidelines (e.g. CONSORT for trials or STROBE for observational studies) are not yet available for Mendelian randomization. However, the MR community has constructed a concise checklist developed to help researchers report their MR findings as clearly as possible: STROBE-MR (Strengthening the Reporting of Observational Studies in Epidemiology using Mendelian Randomization). This checklist was followed where possible (e.g. recommendations for individual level MR were not relevant) and can be found here: https://www.strobe-mr.org/

#### Pre-registration deviations

We have made some minor revisions to our pre-registration (https://osf.io/qfcz4/). In our pre-registration, we included MR-Lap^28^ as one of the sensitivity analyses as this method accounts for sample overlap, which would allow us to include larger samples in our MR analyses. However, we later realized that our instruments employed in the main analyses consisted solely of UK-Biobank, and so did some of the outcomes for which we had planned to use MR-Lap. Also, in some cases, larger sex-stratified samples including UK-Biobank were not available. Therefore, we omitted this sensitivity method. We also included the method MR-RAPS^29^ as a sensitivity analysis in the pre-registration in case we had to use instruments with a lower p-value threshold, however, this turned out to be unnecessary. Lastly, we had planned to use MVMR to investigate the combined effects of several sex hormone traits, contingent on finding significant effects. However, since no significant effects were observed, we omitted this from our methods section.

### Supplementary results

| Genetic correlations  **Table S2.** Genetic correlations performed with LDSC of all included sex hormone traits and severe mental illnesses | | | | |  |
| --- | --- | --- | --- | --- | --- |
| **Sex hormone traits** | **Severe mental illness** | **rg** | **SE** | **p** | **p_FDR_** |
| ***Women only*** |  |  |  |  |  |
| Oestrogen pre-menopause | Depressive disorder | -0.12 | 0.09 | 0.154 | 0.318 |
| Oestrogen pre-menopause | Bipolar disorder | 0.08 | 0.10 | 0.452 | 0.653 |
| Oestrogen pre-menopause | Schizophrenia | 0.13 | 0.08 | 0.112 | 0.284 |
| **Oestrogen post-menopause** | **Depressive disorder** | **-0.22** | **0.07** | **0.002** | **0.020** |
| Oestrogen post-menopause | Bipolar disorder | -0.18 | 0.08 | 0.018 | 0.074 |
| Oestrogen post-menopause | Schizophrenia | -2.6E-03 | 0.06 | 0.966 | 0.993 |
| Oestrogen pre- & post-menopause | Depressive disorder | -4.8E-03 | 0.07 | 0.945 | 0.993 |
| Oestrogen pre- & post-menopause | Bipolar disorder | -7E-04 | 0.08 | 0.993 | 0.993 |
| Oestrogen pre- & post-menopause | Schizophrenia | 0.08 | 0.05 | 0.129 | 0.303 |
| Testosterone pre-menopause | Depressive disorder | -0.05 | 0.05 | 0.374 | 0.609 |
| Testosterone pre-menopause | Bipolar disorder | -0.07 | 0.07 | 0.277 | 0.481 |
| **Testosterone pre-menopause** | **Schizophrenia** | **0.13** | **0.05** | **0.006** | **0.035** |
| Testosterone post-menopause | Depressive disorder | -0.04 | 0.04 | 0.388 | 0.609 |
| Testosterone post-menopause | Bipolar disorder | -0.04 | 0.05 | 0.455 | 0.653 |
| Testosterone post-menopause | Schizophrenia | -2.5E-03 | 0.04 | 0.952 | 0.993 |
| Testosterone pre- & post-menopause | Depressive disorder | -0.05 | 0.03 | 0.053 | 0.147 |
| Testosterone pre- & post-menopause | Bipolar disorder | 0.01 | 0.04 | 0.760 | 0.928 |
| **Testosterone pre- & post-menopause** | **Schizophrenia** | **0.08** | **0.02** | **0.001** | **0.013** |
| SHBG pre-menopause | Depressive disorder | -0.11 | 0.05 | 0.023 | 0.084 |
| SHBG pre-menopause | Bipolar disorder | 0.06 | 0.05 | 0.213 | 0.413 |
| SHBG pre-menopause | Schizophrenia | 0.09 | 0.04 | 0.026 | 0.086 |
| **SHBG post-menopause** | **Depressive disorder** | **-0.13** | **0.04** | **3E-04** | **0.005** |
| SHBG post-menopause | Bipolar disorder | 0.01 | 0.04 | 0.889 | 0.993 |
| SHBG post-menopause | Schizophrenia | 0.07 | 0.03 | 0.030 | 0.089 |
| **SHBG pre- & post-menopause** | **Depressive disorder** | **-0.12** | **0.03** | **1E-04** | **0.003** |
| SHBG pre- & post-menopause | Bipolar disorder | 0.02 | 0.03 | 0.530 | 0.723 |
| **SHBG pre- & post-menopause** | **Schizophrenia** | **0.07** | **0.03** | **0.006** | **0.035** |
| Age of menopause | Depressive disorder | -0.07 | 0.03 | 0.016 | 0.074 |
| Age of menopause | Bipolar disorder | -0.02 | 0.03 | 0.622 | 0.789 |
| Age of menopause | Schizophrenia | -0.03 | 0.03 | 0.234 | 0.428 |
| Age of menarche | Depressive disorder | -0.04 | 0.03 | 0.141 | 0.309 |
| Age of menarche | Bipolar disorder | 2.3E-03 | 0.03 | 0.940 | 0.993 |
| Age of menarche | Schizophrenia | 0.02 | 0.03 | 0.548 | 0.723 |
| ***Men only*** |  |  |  |  |  |
| **Oestrogen** | **Depressive disorder** | **-0.16** | **0.06** | **0.007** | **0.025** |
| Oestrogen | Bipolar disorder | -0.06 | 0.06 | 0.312 | 0.401 |
| Oestrogen | Schizophrenia | -0.03 | 0.05 | 0.544 | 0.612 |
| Testosterone | Depressive disorder | -0.03 | 0.03 | 0.309 | 0.401 |
| Testosterone | Bipolar disorder | 0.04 | 0.04 | 0.275 | 0.401 |
| **Testosterone** | **Schizophrenia** | **0.06** | **0.02** | **0.008** | **0.025** |
| **SHBG** | **Depressive disorder** | **-0.07** | **0.03** | **0.016** | **0.037** |
| SHBG | Bipolar disorder | 0.01 | 0.04 | 0.768 | 0.768 |
| **SHBG** | **Schizophrenia** | **0.08** | **0.02** | **0.001** | **0.012** |
| ***Sexes combined*** |  |  |  |  |  |
| Oestrogen (women) | Depressive disorder | -0.03 | 0.05 | 0.447 | 0.536 |
| Oestrogen (women) | Bipolar disorder | -0.05 | 0.05 | 0.311 | 0.533 |
| Oestrogen (women) | Schizophrenia | 0.04 | 0.04 | 0.375 | 0.536 |
| Oestrogen (men) | Depressive disorder | -0.07 | 0.04 | 0.069 | 0.167 |
| Oestrogen (men) | Bipolar disorder | -0.01 | 0.04 | 0.767 | 0.767 |
| Oestrogen (men) | Schizophrenia | -0.03 | 0.05 | 0.446 | 0.536 |
| **Testosterone** | **Depressive disorder** | **-0.06** | **0.02** | **0.003** | **0.010** |
| Testosterone | Bipolar disorder | 0.01 | 0.02 | 0.610 | 0.665 |
| **Testosterone** | **Schizophrenia** | **0.08** | **0.02** | **3E-05** | **2.4E-04** |
| **SHBG** | **Depressive disorder** | **-0.06** | **0.02** | **1E-04** | **4E-04** |
| SHBG | Bipolar disorder | 0.03 | 0.02 | 0.196 | 0.392 |
| **SHBG** | **Schizophrenia** | **0.08** | **0.02** | **4E-05** | **2.4E-04** |

#### Univariable MR analyses

**Table S3.** Univariable bi-directional MR analysis of the effect of oestrogen/testosterone levels on SMI risk

| **Exposure** | **Outcome** | **SNPs** | **Inverse variance weighted**  **/ Wald ratio** | | | **Weighted median** | | | **Weighted mode** | | | **MR Egger** | | | **IVW outlier corrected**  **MR PRESSO** | | |
| --- | --- | --- | --- | --- | --- | --- | --- | --- | --- | --- | --- | --- | --- | --- | --- | --- | --- |
| *Direction: sex hormone levels to SMI* | | N | OR | 95%CIs | pval | OR | 95%CIs | pval | OR | 95%CIs | pval | OR | 95%CIs | pval | OR | 95%CIs | pval |
| ***Women only*** |  |  |  |  |  |  |  |  |  |  |  |  |  |  |  |  |  |
| **Oestrogen** pre-menopause | Depressive disorder | 2 | 0.98 | 0.42 to 2.26 | 0.954 | - | - | - | - | - | - | - | - | - | - | - | - |
| **Oestrogen** post-menopause | Depressive disorder | 3 | 0.85 | 0.46 to 1.56 | 0.597 | 0.70 | 0.36 to 1.36 | 0.292 | 0.69 | 0.33 to 1.42 | 0.416 | 0.51 | 0.16 to 1.67 | 0.467 | - | - | - |
| **Oestrogen** overall | Depressive disorder | 4 | 1.07 | 0.45 to 2.53 | 0.873 | 1.25 | 0.46 to 3.38 | 0.661 | 1.11 | 0.40 to 3.06 | 0.859 | 0.84 | 0.14 to 5.24 | 0.872 | - | - | - |
| **Oestrogen** pre-menopause | Bipolar disorder | 2 | 0.96 | 0.41 to 2.26 | 0.929 | - | - | - | - | - | - | - | - | - | - | - | - |
| **Oestrogen** post-menopause | Bipolar disorder | 3 | 0.67 | 0.25 to 1.82 | 0.436 | 0.93 | 0.32 to 2.67 | 0.886 | 0.97 | 0.29 to 3.25 | 0.963 | 1.11 | 0.06 to 19.14 | 0.953 | - | - | - |
| **Oestrogen** overall | Bipolar disorder | 5 | 0.47 | 0.16 to 1.41 | 0.180 | 0.73 | 0.19 to 2.80 | 0.651 | 0.80 | 0.21 to 3.02 | 0.763 | 1.94 | 0.05 to 69.14 | 0.741 | - | - | - |
| **Oestrogen** pre-menopause | Schizophrenia | 3 | 0.93 | 0.52 to 1.66 | 0.806 | 1.11 | 0.67 to 1.83 | 0.685 | 1.19 | 0.69 to 2.04 | 0.598 | 1.99 | 0.87 to 4.54 | 0.350 | - | - | - |
| **Oestrogen** post-menopause | Schizophrenia | 4 | 1.10 | 0.61 to 1.96 | 0.760 | 1.20 | 0.76 to 1.91 | 0.440 | 1.23 | 0.73 to 2.09 | 0.492 | 1.54 | 0.47 to 5.07 | 0.551 | - | - | - |
| **Oestrogen** overall | Schizophrenia | 8 | 1.33 | 0.68 to 2.59 | 0.408 | 1.24 | 0.65 to 2.37 | 0.521 | 1.25 | 0.51 to 3.05 | 0.645 | 1.13 | 0.20 to 6.49 | 0.895 | - | - | - |
| **Testosterone** post-menopause | Depressive disorder | 1 | 0.82 | 0.27 to 2.44 | 0.720 | - | - | - | - | - | - | - | - | - | - | - | - |
| **Testosterone** overall | Depressive disorder | 122 | 1.04 | 0.91 to 1.18 | 0.586 | 1.05 | 0.86 to 1.28 | 0.638 | 1.01 | 0.85 to 1.20 | 0.895 | 1.02 | 0.83 to 1.26 | 0.848 | - | - | - |
| **Testosterone** post-menopause | Bipolar disorder | 1 | 1.50 | 0.38 to 6.01 | 0.564 | - | - | - | - | - | - | - | - | - | - | - | - |
| **Testosterone** overall | Bipolar disorder | 128 | 1.07 | 0.92 to 1.23 | 0.384 | 1.32 | 1.05 to 1.66 | 0.017 | 1.31 | 1.02 to 1.67 | 0.034 | 1.17 | 0.92 to 1.50 | 0.202 | - | - | - |
| **Testosterone** post-menopause | Schizophrenia | 1 | 2.16 | 0.80 to 5.87 | 0.131 | - | - | - | - | - | - | - | - | - | - | - | - |
| **Testosterone** overall | Schizophrenia | 178 | 0.996 | 0.90 to 1.10 | 0.938 | 1.04 | 0.90 to 1.21 | 0.572 | 1.07 | 0.89 to 1.27 | 0.478 | 0.99 | 0.82 to 1.19 | 0.882 | - | - | - |
| ***Men only*** |  |  |  |  |  |  |  |  |  |  |  |  |  |  |  |  |  |
| **Oestrogen** | Depressive disorder | 7 | 1.42 | 0.19 to 10.79 | 0.735 | 1.89 | 0.22 to 16.20 | 0.563 | 2.66 | 0.28 to 25.76 | 0.430 | 8.17 | 0.06 to 1058.23 | 0.436 | - | - | - |
| **Oestrogen** | Bipolar disorder | 8 | 0.44 | 0.02 to 9.75 | 0.601 | 0.53 | 0.04 to 6.28 | 0.613 | 0.64 | 0.04 to 10.48 | 0.764 | 10.86 | 0.01 to 19296.41 | 0.555 | - | - | - |
| **Oestrogen** | Schizophrenia | 13 | 1.03 | 0.32 to 3.28 | 0.964 | 2.27 | 0.61 to 8.49 | 0.223 | 4.48 | 1.01 to 19.92 | 0.073 | 0.52 | 0.04 to 7.56 | 0.641 | - | - | - |
| **Testosterone** | Depressive disorder | 99 | 0.96 | 0.80 to 1.14 | 0.632 | 0.88 | 0.67 to 1.15 | 0.339 | 0.88 | 0.63 to 1.21 | 0.423 | 0.94 | 0.64 to 1.37 | 0.735 | - | - | - |
| **Testosterone** | Bipolar disorder | 100 | 1.05 | 0.88 to 1.26 | 0.591 | 0.96 | 0.69 to 1.32 | 0.788 | 1.06 | 0.77 to 1.47 | 0.718 | 0.95 | 0.68 to 1.32 | 0.770 | - | - | - |
| **Testosterone** | Schizophrenia | 147 | **1.09** | **0.98 to 1.20** | **0.099** | 1.13 | 1.001 to 1.28 | 0.048 | 1.13 | 1.002 to 1.28 | 0.047 | 1.06 | 0.90 to 1.26 | 0.486 | 1.09 | 0.998 to 1.19 | 0.057 |
| ***Sexes combined*** |  |  |  |  |  |  |  |  |  |  |  |  |  |  |  |  |  |
| **Testosterone** | Depressive disorder | 166 | 0.99 | 0.87 to 1.12 | 0.825 | 1.11 | 0.88 to 1.39 | 0.384 | 1.07 | 0.86 to 1.33 | 0.539 | 0.96 | 0.77 to 1.21 | 0.734 | - | - | - |
| **Testosterone** | Bipolar disorder | 159 | 1.07 | 0.89 to 1.28 | 0.494 | 0.90 | 0.71 to 1.14 | 0.383 | 0.96 | 0.69 to 1.34 | 0.828 | 1.01 | 0.72 to 1.41 | 0.954 | - | - | - |
| **Testosterone** | Schizophrenia | 159 | 1.09 | 0.89 to 1.33 | 0.410 | 1.14 | 0.91 to 1.43 | 0.240 | 1.15 | 0.92 to 1.43 | 0.228 | 1.003 | 0.69 to 1.45 | 0.989 | - | - | - |
| *Direction: mental illness to sex hormone levels* | | | | | | | | | | | | | | | |  |  |
| Binary outcomes |  | N | OR | 95%CIs | pval | OR | 95%CIs | pval | OR | 95%CIs | pval | OR | 95%CIs | pval | OR | 95%CIs | pval |
| ***Women only*** |  |  |  |  |  |  |  |  |  |  |  |  |  |  |  |  |  |
| Schizophrenia | **Oestrogen** pre-menopause | 18 | 0.99 | 0.96 to 1.02 | 0.538 | 0.999 | 0.95 to 1.05 | 0.958 | 1.04 | 0.95 to 1.14 | 0.415 | - | - | - | - | - | - |
| Schizophrenia | **Oestrogen** post-menopause | 18 | 0.99 | 0.96 to 1.01 | 0.340 | 0.99 | 0.96 to 1.03 | 0.743 | 0.98 | 0.93 to 1.05 | 0.611 | - | - | - | - | - | - |
| Schizophrenia | **Oestrogen** overall | 18 | 1.00006 | 0.98 to 1.02 | 0.995 | 1.001 | 0.98 to 1.03 | 0.961 | 1.01 | 0.96 to 1.06 | 0.755 | - | - | - | - | - | - |
| ***Men only*** |  |  |  |  |  |  |  |  |  |  |  |  |  |  |  |  |  |
| Schizophrenia | **Oestrogen** |  |  |  |  |  |  |  |  |  |  | - | - | - | - |  |  |
| Continuous outcomes | | N | beta | 95%CIs | pval | beta | 95%CIs | pval | beta | 95%CIs | pval | beta | 95%CIs | pval | beta | 95%CIs | pval |
| ***Women only*** |  |  |  |  |  |  |  |  |  |  |  |  |  |  |  |  |  |
| Schizophrenia | **Testosterone** pre-menopause | 18 | -0.01 | -0.06 to 1.28 | 0.6398 | -0.01 | -0.07 to 1.32 | 0.659 | -0.042 | -0.15 to 1.00 | 0.481 | - | - | - | - | - | - |
| Schizophrenia | **Testosterone** post-menopause | 18 | 1.9E-03 | -0.03 to 0.03 | 0.898 | -0.01 | -0.05 to 0.03 | 0.721 | -0.02 | -0.08 to 0.05 | 0.606 |  |  |  |  |  |  |
| Schizophrenia | **Testosterone** overall | 22 | -1.4E-03 | -0.03 to 0.02 | 0.909 | -0.02 | -0.04 to 0.01 | 0.138 | -0.03 | -0.06 to -0.003 | 0.043 | - | - | - | - | - | - |
| ***Men only*** |  |  |  |  |  |  |  |  |  |  |  |  |  |  |  |  |  |
| Schizophrenia | **Testosterone** | 53 | -3.7E-04 | -0.03 to 0.02 | 0.977 | 0.01 | -0.01 to 0.03 | 0.423 | 0.05 | 3.6E-03 to 0.1 | 0.040 | - | - | - | - | - | - |
| ***Sexes combined*** |  |  |  |  |  |  |  |  |  |  |  |  |  |  |  |  |  |
| Depressive disorder | **Testosterone** | 2 | 5.7E-04 | -0.03 to 0.03 | 0.973 | - | - | - | - | - | - | - | - | - | - | - | - |
| Bipolar disorder | **Testosterone** | 49 | -3.0E-03 | -0.01 to 0.01 | 0.574 | 2.5E-03 | -0.01 to 0.01 | 0.624 | 0.01 | -0.01 to 0.04 | 0.245 | - | - | - | - | - | - |
| Schizophrenia | **Testosterone** | 177 | **0.01** | **-5.4E-04 to 0.01** | **0.076** | 4.8E-03 | -6.5E-04 to 0.01 | 0.084 | 2.6E-03 | -0.01 to 0.02 | 0.711 | -1.5E-03 | -0.02 to 0.02 | 0.901 | 0.01 | 1.1E-03 to 0.01 | 0.017 |

**Table S4.** Univariable bi-directional MR analysis of the effect of other sex hormone traits on SMI risk (exploratory analyses)

| **Exposure** | **Outcome** | **SNPs** | **Inverse variance weighted** | | | | **Weighted median** | | | | | | | **Weighted mode** | | | | | | **MR Egger** | | | | | | | **IVW outlier corrected (MR PRESSO)** | | | |  |  |
| --- | --- | --- | --- | --- | --- | --- | --- | --- | --- | --- | --- | --- | --- | --- | --- | --- | --- | --- | --- | --- | --- | --- | --- | --- | --- | --- | --- | --- | --- | --- | --- | --- |
| Direction: sex hormone traits to SMI | | N | OR | 95%CI | pval | OR | | | | 95%CI | | pval | OR | | | 95%CI | | | pval | | OR | 95%CI | | pval | | OR | | 95%CI | | | | pval |
| **WOMEN** |  |  |  |  |  | | |  |  | |  | | | |  | |  |  | |  | | |  | |  | |  | |  |  |  |  |
| **SHBG** pre-menopause | Depressive disorder | 16 | **1.16** | **1.001 to 1.35** | **0.048** | | | 1.16 | 0.96 to 1.42 | | 0.125 | | | | 1.13 | | 0.89 to 1.43 | 0.332 | | 1.15 | | | 0.79 to 1.69 | | 0.477 | | - | | - | - |  |  |
| **SHBG** post-menopause | Depressive disorder | 53 | **1.24** | **1.10 to 1.40** | **0.001** | | | 1.18 | 0.99 to 1.41 | | 0.069 | | | | 1.15 | | 0.93 to 1.41 | 0.200 | | 1.06 | | | 0.80 to 1.40 | | 0.690 | | - | | - | - |  |  |
| **SHBG** pre-menopause | Bipolar disorder | 15 | 0.90 | 0.71 to 1.14 | 0.374 | | | 0.93 | 0.71 to 1.21 | | 0.586 | | | | 0.95 | | 0.64 to 1.43 | 0.818 | | 1.06 | | | 0.56 to 2.01 | | 0.858 | | - | | - | - |  |  |
| **SHBG** post-menopause | Bipolar disorder | 54 | 0.96 | 0.81 to 1.14 | 0.659 | | | 1.03 | 0.81 to 1.30 | | 0.826 | | | | 1.03 | | 0.70 to 1.50 | 0.894 | | 1.24 | | | 0.82 to 1.85 | | 0.310 | | - | | - | - |  |  |
| **SHBG** pre-menopause | Schizophrenia | 22 | 1.07 | 0.93 to 1.23 | 0.349 | | | 0.97 | 0.84 to 1.13 | | 0.724 | | | | 0.93 | | 0.75 to 1.14 | 0.474 | | 1.26 | | | 0.88 to 1.81 | | 0.222 | | - | | - | - |  |  |
| **SHBG** post-menopause | Schizophrenia | 82 | 1.03 | 0.92 to 1.15 | 0.580 | | | 0.99 | 0.87 to 1.13 | | 0.929 | | | | 0.93 | | 0.77 to 1.13 | 0.488 | | 1.14 | | | 0.88 to 1.45 | | 0.323 | | - | | - | - |  |  |
| **Progesterone** | Depressive disorder | 2 | 1.02 | 0.86 to 1.21 | 0.796 | | | - | - | | - | | | | - | | - | - | | - | | | - | | - | | - | | - | - |  |  |
| **Progesterone** | Bipolar disorder | 2 | 1.12 | 0.89 to 1.42 | 0.323 | | | - | - | | - | | | | - | | - | - | | - | | | - | | - | | - | | - | - |  |  |
| **Progesterone** | Schizophrenia | 2 | 1.09 | 0.94 to 1.28 | 0.252 | | | - | - | | - | | | | - | | - | - | | - | | | - | | - | | - | | - | - |  |  |
| **Age at menarche** | Depressive disorder | 245 | 0.94 | 0.86 to 1.02 | 0.143 | | | 0.92 | 0.80 to 1.06 | | 0.265 | | | | 0.91 | | 0.73 to 1.14 | 0.432 | | 2.39 | | | 1.54 to 3.70 | | 1.7E-04 | | - | | - | - |  |  |
| **Age at menarche** | Bipolar disorder | 248 | 1.002 | 0.89 to 1.12 | 0.973 | | | 0.92 | 0.78 to 1.09 | | 0.346 | | | | 0.89 | | 0.64 to 1.24 | 0.496 | | 1.14 | | | 0.62 to 2.11 | | 0.675 | | - | | - | - |  |  |
| **Age at menarche** | Schizophrenia | 316 | 1.04 | 0.95 to 1.14 | 0.426 | | | 1.008 | 0.91 to 1.12 | | 0.879 | | | | 1.02 | | 0.69 to 1.51 | 0.910 | | 1.46 | | | 0.92 to 2.31 | | 0.112 | | - | | - | - |  |  |
| **Age at menopause** | Depressive disorder | 156 | 0.99 | 0.96 to 1.02 | 0.375 | | | 0.96 | 0.91 to 1.01 | | 0.121 | | | | 0.96 | | 0.90 to 1.02 | 0.168 | | 0.97 | | | 0.91 to 1.04 | | 0.430 | | - | | - | - |  |  |
| **Age at menopause** | Bipolar disorder | 163 | 1.02 | 0.97 to 1.07 | 0.473 | | | 1.04 | 0.98 to 1.10 | | 0.245 | | | | 1.04 | | 0.96 to 1.12 | 0.309 | | 1.02 | | | 0.90 to 1.15 | | 0.792 | | - | | - | - |  |  |
| **Age at menopause** | Schizophrenia | 216 | 1.01 | 0.98 to 1.04 | 0.612 | | | 1.004 | 0.97 to 1.04 | | 0.834 | | | | 0.999 | | 0.96 to 1.04 | 0.958 | | ###### | | | 0.93 to 1.08 | | 0.99995 | | - | | - | - |  |  |
| **MEN** |  |  |  |  |  | | |  |  | |  | | | |  | |  |  | |  | | |  | |  | |  | |  |  |  |  |
| **SHBG** | Depressive disorder | 123 | 1.26 | 0.93 to 1.72 | 0.136 | | | 0.90 | 0.55 to 1.47 | | 0.675 | | | | 0.91 | | 0.56 to 1.45 | 0.683 | | 0.77 | | | 0.44 to 1.34 | | 0.355 | | - | | - | - |  |  |
| **SHBG** | Bipolar disorder | 125 | 0.999 | 0.68 to 1.48 | 0.995 | | | 0.89 | 0.51 to 1.53 | | 0.664 | | | | 0.89 | | 0.44 to 1.81 | 0.751 | | 1.01 | | | 0.49 to 2.11 | | 0.973 | | - | | - | - |  |  |
| **SHBG** | Schizophrenia | 177 | 1.03 | 0.83 to 1.27 | 0.815 | | | 1.08 | 0.83 to 1.42 | | 0.549 | | | | 1.07 | | 0.73 to 1.59 | 0.721 | | 1.13 | | | 0.74 to 1.71 | | 0.575 | | - | | - | - |  |  |
| **SEXES COMBINED** |  |  |  |  |  | | |  |  | |  | | | |  | |  |  | |  | | |  | |  | |  | |  |  |  |  |
| **SHBG** | Depressive disorder | 335 | 1.05 | 0.94 to 1.18 | 0.386 | | | 1.06 | 0.88 to 1.28 | | 0.410 | | | | 0.95 | | 0.75 to 1.19 | 0.628 | | 0.92 | | | 0.75 to 1.14 | | 0.453 | | - | | - | - |  |  |
| **SHBG** | Bipolar disorder | 307 | 0.97 | 0.81 to 1.15 | 0.692 | | | 0.93 | 0.77 to 1.14 | | 0.506 | | | | 0.95 | | 0.71 to 1.27 | 0.717 | | 0.90 | | | 0.65 to 1.24 | | 0.510 | | - | | - | - |  |  |
| **SHBG** | Schizophrenia | 306 | 1.07 | 0.90 to 1.27 | 0.465 | | | 1.06 | 0.88 to 1.28 | | 0.839 | | | | 1.03 | | 0.77 to 1.37 | 0.846 | | 1.03 | | | 0.75 to 1.43 | | 0.839 | | - | | - | - |  |  |
| Direction: mental illness to sex hormone traits | | N | beta | 95%CI | pval | | | beta | 95%CI | | pval | | | | beta | | 95%CI | pval | | beta | | | 95%CI | | pval | | beta | | 95%CI | pval |  |  |
| **WOMEN** |  |  |  |  |  | | |  |  | |  | | | |  | |  |  | |  | | |  | |  | |  | |  |  |  |  |
| Schizophrenia | **SHBG** pre-menopause | 18 | 0.03 | -0.01 to 0.06 | 0.137 | | | 0.05 | -3.1E-03 to 0.09 | | 0.066 | | | | 0.06 | | -0.03 to 0.14 | 0.225 | | - | | | - | | - | | - | | - | - |  |  |
| Schizophrenia | **SHBG** post-menopause | 18 | **0.05** | **0.02 to 0.09** | **0.002** | | | 0.05 | 0.01 to 0.09 | | 0.007 | | | | 0.07 | | 0.003 to 0.13 | 0.055 | | - | | | - | | - | | 0.05 | | 0.02 to 0.09 | 0.002 |  |  |
| Schizophrenia | **Progesterone** | 22 | **-0.31** | **-0.62 to -4.6E-03** | **0.047** | | | -0.37 | -0.72 to -0.03 | | 0.035 | | | | -0.37 | | -0.69 to -0.05 | 0.033 | | 0.32 | | | -2.03 to 2.66 | | 0.794 | | -0.31 | | -0.62 to -4.6E-03 | 0.047 |  |  |
| Schizophrenia | **Age at menarche** | 10 | 0.01 | -0.04 to 0.05 | 0.828 | | | 0.01 | -0.05 to 0.06 | | 0.867 | | | | -0.06 | | -0.19 to 0.06 | 0.360 | | - | | | - | | - | | - | | - | - |  |  |
| Schizophrenia | **Age at menopause** | 10 | 0.02 | -0.12 to 0.16 | 0.766 | | | 0.02 | -0.17 to 0.21 | | 0.821 | | | | -0.16 | | -0.49 to 0.17 | 0.374 | | - | | | - | | - | | - | | - | - |  |  |
| **MEN** |  |  |  |  |  | | |  |  | |  | | | |  | |  |  | |  | | |  | |  | |  | |  |  |  |  |
| Schizophrenia | **SHBG** | 53 | 3.0E-03 | -0.01 to 0.01 | 0.472 | | | 4.4E-03 | -2.6E-03 to 0.01 | | 0.215 | | | | 0.01 | | -4.7E-03 to 0.02 | 0.267 | | - | | | - | | - | | - | |  |  |  |  |
| **SEXES COMBINED** |  |  |  |  |  | | |  |  | |  | | | |  | |  |  | |  | | |  | |  | |  | |  |  |  |  |
| Depressive disorder | **SHBG** | 2 | -0.03 | -0.07 to 0.01 | 0.112 | | | - | - | | - | | | | - | | - | - | | - | | | - | | - | | - | | - | - |  |  |
| Bipolar disorder | **SHBG** | 49 | **0.01** | **-1.1E-03 to 0.02** | **0.074** | | | 0.02 | 0.01 to 0.02 | | 1E-05 | | | | 0.02 | | 0.01 to 0.03 | 0.007 | | - | | | - | | - | | 0.02 | | 8.4E-03 to 0.03 | 8.2E-05 |  |  |
| Schizophrenia | **SHBG** | 177 | **0.01** | **3.8E-03 to 0.01** | **0.001** | | | 0.01 | 3.0E-03 to 0.01 | | 0.001 | | | | 0.01 | | -9.2E-04 to 0.02 | 0.073 | | -0.03 | | | -0.05 to -0.01 | | 0.012 | | 0.01 | | 3.6E-03 to 0.01 | 2.4E-04 |  |  |

**Table S5.** Generalized summary data-based Mendelian randomization (GSMR) analysis for all univariable MR analyses (main + exploratory analysis)

| **Exposure** | **Outcome** | **beta** | **se** | **pval** |
| --- | --- | --- | --- | --- |
| Testosterone [Men only] | Schizophrenia [Men only] | 0.12 | 0.04 | 0.001 |
| SHBG pre-menopause [Women only] | Depressive disorder [Women only] | 0.14 | 0.09 | 0.098 |
| SHBG post-menopause [Women only] | Depressive disorder [Women only] | 0.22 | 0.07 | 0.001 |
| Schizophrenia [Sexes combined] | Testosterone [Sexes combined] | 4.0E-03 | 1.8E-03 | 0.022 |
| Schizophrenia [Women only] | SHBG post-menopause | 0.05 | 0.01 | 5.0E-05 |
| Schizophrenia [Women only] | Progesterone | -0.31 | 0.16 | 0.047 |
| Bipolar disorder [Sexes combined] | SHBG [Sexes combined] | 0.01 | 1.3E-03 | 2.2E-11 |
| Schizophrenia [Sexes combined] | SHBG [Sexes combined] | 0.02 | 2.5E-03 | 2.2E-18 |

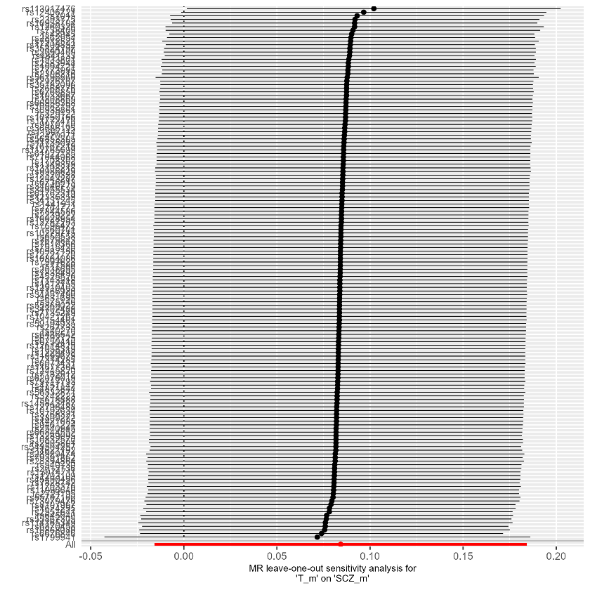
  **Figure S3.** Leave-one-out analysis of MR IVW regression analysis to investigate the effect of testosterone levels on schizophrenia in men

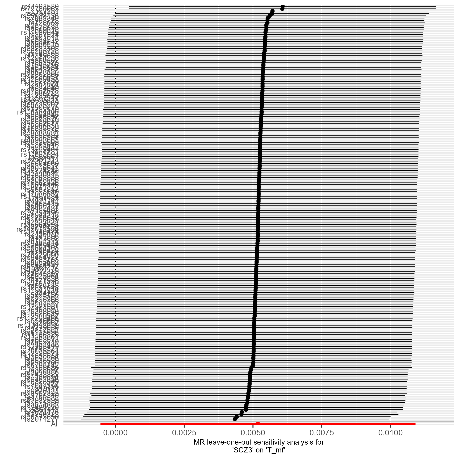
  **Figure S4.** Leave-one-out analysis of MR IVW regression analysis to investigate the effect of liability to schizophrenia on testosterone levels in sexes combined

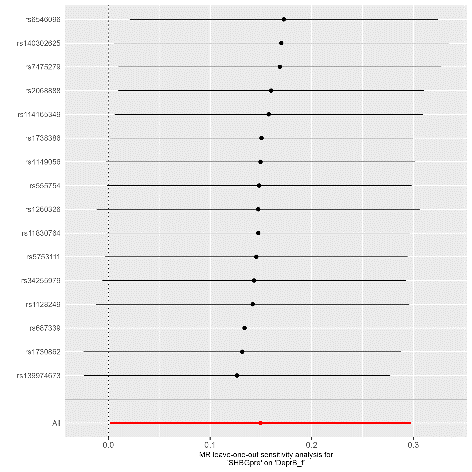
 **Figure S5.** Leave-one-out analysis of MR IVW regression analysis to investigate the effect of pre-menopausal SHBG levels on depressive disorder in women

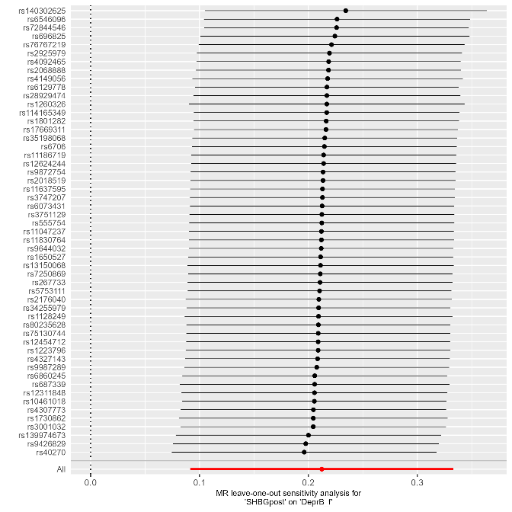
 **Figure S6.** Leave-one-out analysis of MR IVW regression analysis to investigate the effect of post-menopausal SHBG levels on depressive disorder in women

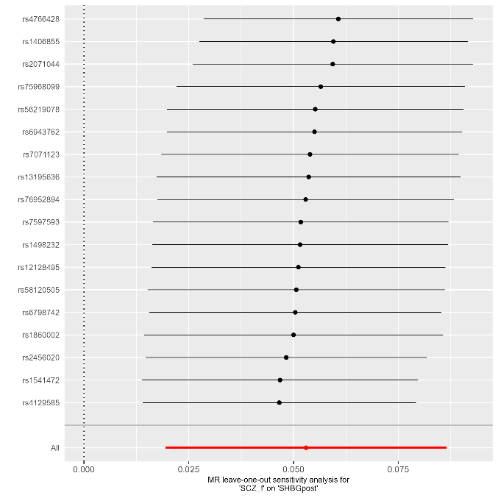
 **Figure S7.** Leave-one-out analysis of MR IVW regression analysis to investigate the effect of liability to schizophrenia on post-menopausal SHBG levels in women

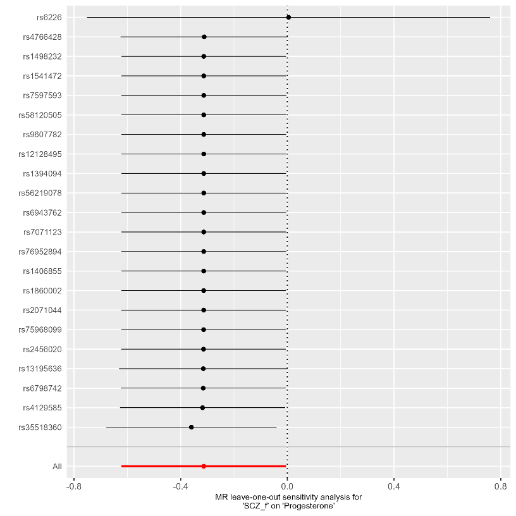
  **Figure S8.** Leave-one-out analysis of MR IVW regression analysis to investigate the effect of liability to schizophrenia on progesterone levels in women

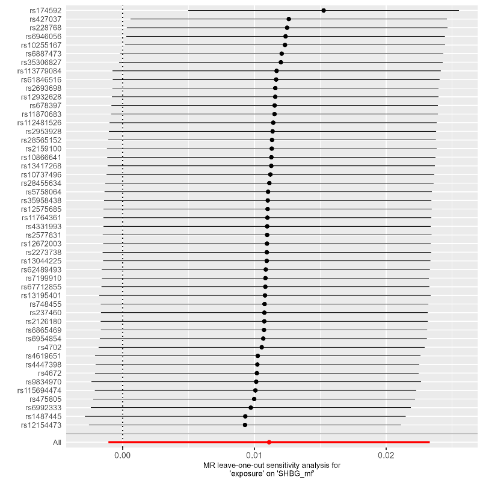
 **Figure S9.** Leave-one-out analysis of MR IVW regression analysis to investigate the effect of liability to bipolar disorder on SHBG levels in sexes combined

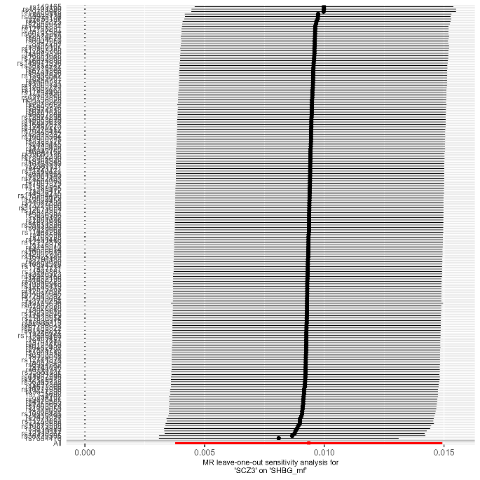
 **Figure S10.** Leave-one-out analysis of MR IVW regression analysis to investigate the effect of liability to schizophrenia on SHBG levels in sexes combined

**Table S6.** Univariable MR analyses after excluding SNPs with Steiger filtering (without a p-value threshold) for main + exploratory analyses

| **Exposure** | **Outcome** | **Analysis** | **N SNPs in original analysis** | **N SNPs in Steiger analysis** | **Method** | **beta** | **se** | **pval** |
| --- | --- | --- | --- | --- | --- | --- | --- | --- |
| Testosterone [overall] | Schizophrenia | Men only | 147 | 145 | IVW | 0.084 | 0.049 | 0.087 |
| Testosterone [overall] | Schizophrenia | Men only | 147 | 145 | Weighted median | 0.123 | 0.066 | 0.064 |
| Testosterone [overall] | Schizophrenia | Men only | 147 | 145 | Weighted mode | 0.121 | 0.061 | 0.050 |
| Testosterone [overall] | Schizophrenia | Men only | 147 | 145 | MR-Egger | 0.051 | 0.082 | 0.533 |
| SHBG [pre-menopause] | Schizophrenia | Women only | 16 | 16 | IVW | - | - | - |
| SHBG [pre-menopause] | Schizophrenia | Women only | 16 | 16 | Weighted median | - | - | - |
| SHBG [pre-menopause] | Schizophrenia | Women only | 16 | 16 | Weighted mode | - | - | - |
| SHBG [pre-menopause] | Schizophrenia | Women only | 16 | 16 | MR-Egger | - | - | - |
| SHBG [post-menopause] | Schizophrenia | Women only | 53 | 53 | IVW | - | - | - |
| SHBG [post-menopause] | Schizophrenia | Women only | 53 | 53 | Weighted median | - | - | - |
| SHBG [post-menopause] | Schizophrenia | Women only | 53 | 53 | Weighted mode | - | - | - |
| SHBG [post-menopause] | Schizophrenia | Women only | 53 | 53 | MR-Egger | - | - | - |
| Schizophrenia | Testosterone | Sexes combined | 177 | 177 | IVW | - | - | - |
| Schizophrenia | Testosterone | Sexes combined | 177 | 177 | Weighted median | - | - | - |
| Schizophrenia | Testosterone | Sexes combined | 177 | 177 | Weighted mode | - | - | - |
| Schizophrenia | Testosterone | Sexes combined | 177 | 177 | MR-Egger | - | - | - |
| Schizophrenia | SHBG | Sexes combined | 177 | 176 | IVW | 0.008 | 0.003 | 0.002 |
| Schizophrenia | SHBG | Sexes combined | 177 | 176 | Weighted median | 0.007 | 0.002 | 0.001 |
| Schizophrenia | SHBG | Sexes combined | 177 | 176 | Weighted mode | 0.010 | 0.006 | 0.097 |
| Schizophrenia | SHBG | Sexes combined | 177 | 176 | MR-Egger | -0.003 | 0.010 | 0.737 |
| Bipolar disorder | SHBG | Sexes combined | 49 | 49 | IVW | - | - | - |
| Bipolar disorder | SHBG | Sexes combined | 49 | 49 | Weighted median | - | - | - |
| Bipolar disorder | SHBG | Sexes combined | 49 | 49 | Weighted mode | - | - | - |
| Bipolar disorder | SHBG | Sexes combined | 49 | 49 | MR-Egger | - | - | - |
| Schizophrenia | SHBG [post-menopause] | Women only | 18 | 18 | IVW | - | - | - |
| Schizophrenia | SHBG [post-menopause] | Women only | 18 | 18 | Weighted median | - | - | - |
| Schizophrenia | SHBG [post-menopause] | Women only | 18 | 18 | Weighted mode | - | - | - |
| Schizophrenia | SHBG [post-menopause] | Women only | 18 | 18 | MR-Egger | - | - | - |
| Schizophrenia | Progesterone | Women only | 22 | 21 | IVW | 0.005 | 0.386 | 0.990 |
| Schizophrenia | Progesterone | Women only | 22 | 21 | Weighted median | -0.081 | 0.462 | 0.861 |
| Schizophrenia | Progesterone | Women only | 22 | 21 | Weighted mode | 0.333 | 0.402 | 0.417 |
| Schizophrenia | Progesterone | Women only | 22 | 21 | MR-Egger | 0.317 | 1.198 | 0.794 |

**Table S7.** MR-Egger intercepts for all univariable MR analyses, indicating evidence of horizontal pleiotropy

| **Exposure** | **Outcome** | **Analysis** | **beta** | **se** | **pval** |
| --- | --- | --- | --- | --- | --- |
| Oestrogen [pre-menopause] | Depressive disorder | Women only | N.A. | N.A. | N.A. |
| Oestrogen [pre-menopause] | Bipolar disorder | Women only | N.A. | N.A. | N.A. |
| Oestrogen [pre-menopause] | Schizophrenia | Women only | -0.05 | 0.03 | 0.289 |
| Oestrogen [post-menopause] | Bipolar disorder | Women only | -0.02 | 0.05 | 0.763 |
| Oestrogen [post-menopause] | Schizophrenia | Women only | -0.02 | 0.03 | 0.573 |
| Oestrogen [post-menopause] | Depressive disorder | Women only | 0.03 | 0.03 | 0.506 |
| Oestrogen [overall] | Bipolar disorder | Women only | -0.03 | 0.04 | 0.474 |
| Oestrogen [overall] | Schizophrenia | Women only | 4.3E-03 | 0.02 | 0.851 |
| Oestrogen [overall] | Depressive disorder | Women only | 0.03 | 0.02 | 0.403 |
| Testosterone [post-menopause] | Depressive disorder | Women only | N.A. | N.A. | N.A. |
| Testosterone [post-menopause] | Bipolar disorder | Women only | N.A. | N.A. | N.A. |
| Testosterone [post-menopause] | Schizophrenia | Women only | N.A. | N.A. | N.A. |
| Testosterone [overall] | Depressive disorder | Women only | 6.9E-04 | 4.0E-03 | 0.865 |
| Testosterone [overall] | Bipolar disorder | Women only | -4.3E-03 | 4.5E-03 | 0.342 |
| Testosterone [overall] | Schizophrenia | Women only | 3.9E-04 | 3.1E-03 | 0.899 |
| SHBG [pre-menopause] | Depressive disorder | Women only | 5.8E-04 | 0.01 | 0.969 |
| SHBG [pre-menopause] | Bipolar disorder | Women only | -0.01 | 0.02 | 0.593 |
| SHBG [pre-menopause] | Schizophrenia | Women only | -0.01 | 0.01 | 0.341 |
| SHBG [post-menopause] | Depressive disorder | Women only | 0.01 | 0.01 | 0.222 |
| SHBG [post-menopause] | Bipolar disorder | Women only | -0.01 | 0.01 | 0.188 |
| SHBG [post-menopause] | Schizophrenia | Women only | -0.01 | 0.01 | 0.404 |
| Progesterone | Depressive disorder | Women only | N.A. | N.A. | N.A. |
| Progesterone | Bipolar disorder | Women only | N.A. | N.A. | N.A. |
| Progesterone | Schizophrenia | Women only | N.A. | N.A. | N.A. |
| Age of menarche | Depressive disorder | Women only | -0.02 | 0.01 | 0.002 |
| Age of menarche | Bipolar disorder | Women only | -3.5E-03 | 0.01 | 0.655 |
| Age of menarche | Schizophrenia | Women only | -0.01 | 0.01 | 0.084 |
| Age of menopause | Depressive disorder | Women only | 2.8E-03 | 0.01 | 0.661 |
| Age of menopause | Bipolar disorder | Women only | 2.0E-04 | 0.01 | 0.985 |
| Age of menopause | Schizophrenia | Women only | 1.7E-03 | 0.01 | 0.812 |
| Oestrogen | Schizophrenia | Men only | -1.3E-03 | 0.01 | 0.933 |
| Oestrogen | Depressive disorder | Men only | -0.02 | 0.03 | 0.470 |
| Oestrogen | Bipolar disorder | Men only | -0.04 | 0.04 | 0.389 |
| Testosterone | Depressive disorder | Men only | 8.0E-04 | 0.01 | 0.894 |
| Testosterone | Bipolar disorder | Men only | 3.9E-03 | 0.01 | 0.485 |
| Testosterone | Schizophrenia | Men only | 1.1E-03 | 3.0E-03 | 0.721 |
| SHBG | Depressive disorder | Men only | 0.01 | 4.6E-03 | 0.039 |
| SHBG | Bipolar disorder | Men only | -2.6E-04 | 0.01 | 0.964 |
| SHBG | Schizophrenia | Men only | -1.7E-03 | 3.2E-03 | 0.607 |
| Testosterone | Depressive disorder | Sexes combined | 4.4E-04 | 1.7E-03 | 0.794 |
| Testosterone | Bipolar disorder | Sexes combined | 9.1E-04 | 2.4E-03 | 0.709 |
| Testosterone | Schizophrenia | Sexes combined | 1.4E-03 | 2.7E-03 | 0.607 |
| SHBG | Depressive disorder | Sexes combined | 1.9E-03 | 1.3E-03 | 0.147 |
| SHBG | Bipolar disorder | Sexes combined | 1.0E-03 | 1.9E-03 | 0.596 |
| SHBG | Schizophrenia | Sexes combined | 4.4E-04 | 2.0E-03 | 0.825 |
| Schizophrenia | Oestrogen [pre-menopause] | Women only | -0.01 | 0.01 | 0.297 |
| Schizophrenia | Oestrogen [post-menopause] | Women only | 0.01 | 0.01 | 0.479 |
| Schizophrenia | Oestrogen [overall] | Women only | -0.01 | 4.9E-03 | 0.165 |
| Schizophrenia | Testosterone [pre-menopause] | Women only | 0.03 | 0.01 | 0.022 |
| Schizophrenia | Testosterone [post-menopause] | Women only | 1.6E-03 | 0.01 | 0.841 |
| Schizophrenia | Testosterone [overall] | Women only | -2.4E-03 | 0.01 | 0.698 |
| Schizophrenia | SHBG [pre-menopause] | Women only | 0.01 | 0.01 | 0.559 |
| Schizophrenia | SHBG [post-menopause] | Women only | 4.0E-03 | 0.01 | 0.655 |
| Schizophrenia | Progesterone | Women only | -0.08 | 0.14 | 0.601 |
| Schizophrenia | Age of menarche | Women only | 0.02 | 0.01 | 0.085 |
| Schizophrenia | Age of menopause | Women only | -0.05 | 0.03 | 0.157 |
| Schizophrenia | Oestrogen | Men only | -4.6E-04 | 5.3E-04 | 0.392 |
| Schizophrenia | Testosterone | Men only | -2.9E-03 | 4.5E-03 | 0.522 |
| Schizophrenia | SHBG | Men only | -7.3E-04 | 1.5E-03 | 0.621 |
| Depressive disorder | SHBG | Sexes combined | N.A. | N.A. | N.A. |
| Depressive disorder | Testosterone | Sexes combined | N.A. | N.A. | N.A. |
| Schizophrenia | Testosterone | Sexes combined | 4.5E-04 | 7.7E-04 | 0.561 |
| Schizophrenia | SHBG | Sexes combined | 1.0E-03 | 7.5E-04 | 0.183 |
| Bipolar disorder | Testosterone | Sexes combined | -1.2E-03 | 1.8E-03 | 0.507 |
| Bipolar disorder | SHBG | Sexes combined | -4.2E-04 | 2.1E-03 | 0.840 |

**Table S8.** Cochran’s Q-statistic for all univariable MR analyses, indicating evidence for heterogeneity

| **Exposure** | **Outcome** | **Analysis** | **method** | **Q** | **Q_df** | **Q_pval** |
| --- | --- | --- | --- | --- | --- | --- |
| Bipolar disorder | Testosterone | Sexes combined | MR Egger | 141.88 | 47 | 1.9E-11 |
| Bipolar disorder | Testosterone | Sexes combined | IVW | 143.23 | 48 | 2.1E-11 |
| Bipolar disorder | SHBG | Sexes combined | MR Egger | 364.17 | 47 | 1.3E-50 |
| Bipolar disorder | SHBG | Sexes combined | IVW | 364.49 | 48 | 3.1E-50 |
| Depressive disorder | Testosterone | Sexes combined | IVW | 0.21 | 1 | 0.647 |
| Depressive disorder | SHBG | Sexes combined | IVW | 2.67 | 1 | 0.102 |
| Schizophrenia | Testosterone | Sexes combined | MR Egger | 534.14 | 175 | 5.4E-38 |
| Schizophrenia | Testosterone | Sexes combined | IVW | 535.17 | 176 | 6.7E-38 |
| Schizophrenia | SHBG | Sexes combined | MR Egger | 958.66 | 175 | 2.7E-108 |
| Schizophrenia | SHBG | Sexes combined | IVW | 968.44 | 176 | 1.2E-109 |
| Age of menarche | Schizophrenia | Women only | MR Egger | 231.71 | 118 | 2.1E-09 |
| Age of menarche | Schizophrenia | Women only | IVW | 237.68 | 119 | 6.4E-10 |
| Age of menarche | Depressive disorder | Women only | MR Egger | 115.69 | 107 | 0.266 |
| Age of menarche | Depressive disorder | Women only | IVW | 126.36 | 108 | 0.109 |
| Age of menarche | Bipolar disorder | Women only | MR Egger | 133.09 | 108 | 0.051 |
| Age of menarche | Bipolar disorder | Women only | IVW | 133.34 | 109 | 0.057 |
| Age of menopause | Schizophrenia | Women only | MR Egger | 89.83 | 49 | 3.4E-04 |
| Age of menopause | Schizophrenia | Women only | IVW | 89.93 | 50 | 4.6E-04 |
| Age of menopause | Depressive disorder | Women only | MR Egger | 33.45 | 41 | 0.793 |
| Age of menopause | Depressive disorder | Women only | IVW | 33.64 | 42 | 0.818 |
| Age of menopause | Bipolar disorder | Women only | MR Egger | 56.60 | 41 | 0.053 |
| Age of menopause | Bipolar disorder | Women only | IVW | 56.60 | 42 | 0.066 |
| Oestrogen [overall] | Schizophrenia | Women only | MR Egger | 13.35 | 6 | 0.038 |
| Oestrogen [overall] | Schizophrenia | Women only | IVW | 13.44 | 7 | 0.062 |
| Oestrogen [overall] | Schizophrenia | Women only | MR Egger | 13.56 | 6 | 0.035 |
| Oestrogen [overall] | Schizophrenia | Women only | IVW | 13.65 | 7 | 0.058 |
| Oestrogen [overall] | Depressive disorder | Women only | MR Egger | 2.12 | 2 | 0.347 |
| Oestrogen [overall] | Depressive disorder | Women only | IVW | 3.29 | 3 | 0.349 |
| Oestrogen [overall] | Depressive disorder | Women only | MR Egger | 0.49 | 1 | 0.485 |
| Oestrogen [overall] | Depressive disorder | Women only | IVW | 2.47 | 2 | 0.291 |
| Oestrogen [overall] | Bipolar disorder | Women only | MR Egger | 3.12 | 3 | 0.374 |
| Oestrogen [overall] | Bipolar disorder | Women only | IVW | 3.91 | 4 | 0.419 |
| Oestrogen [overall] | Bipolar disorder | Women only | MR Egger | 3.55 | 3 | 0.314 |
| Oestrogen [overall] | Bipolar disorder | Women only | IVW | 4.34 | 4 | 0.362 |
| Oestrogen | Schizophrenia | Men only | MR Egger | 13.21 | 9 | 0.153 |
| Oestrogen | Schizophrenia | Men only | IVW | 13.22 | 10 | 0.211 |
| Oestrogen | Depressive disorder | Men only | MR Egger | 7.08 | 5 | 0.215 |
| Oestrogen | Depressive disorder | Men only | IVW | 7.95 | 6 | 0.242 |
| Oestrogen | Bipolar disorder | Men only | MR Egger | 14.93 | 6 | 0.021 |
| Oestrogen | Bipolar disorder | Men only | IVW | 17.07 | 7 | 0.017 |
| Oestrogen [post-menopause] | Schizophrenia | Women only | MR Egger | 5.30 | 2 | 0.071 |
| Oestrogen [post-menopause] | Schizophrenia | Women only | IVW | 6.48 | 3 | 0.090 |
| Oestrogen [post-menopause] | Depressive disorder | Women only | MR Egger | 1.20 | 1 | 0.273 |
| Oestrogen [post-menopause] | Depressive disorder | Women only | IVW | 2.36 | 2 | 0.308 |
| Oestrogen [post-menopause] | Bipolar disorder | Women only | MR Egger | 2.14 | 1 | 0.143 |
| Oestrogen [post-menopause] | Bipolar disorder | Women only | IVW | 2.47 | 2 | 0.291 |
| Oestrogen [pre-menopause] | Schizophrenia | Women only | MR Egger | 0.14 | 1 | 0.712 |
| Oestrogen [pre-menopause] | Schizophrenia | Women only | IVW | 4.34 | 2 | 0.114 |
| Oestrogen [pre-menopause] | Depressive disorder | Women only | IVW | 2.17 | 1 | 0.141 |
| Oestrogen [pre-menopause] | Bipolar disorder | Women only | IVW | 0.03 | 1 | 0.859 |
| Progesterone | Schizophrenia | Women only | IVW | 0.03 | 1 | 0.870 |
| Progesterone | Depressive disorder | Women only | IVW | 0.53 | 1 | 0.467 |
| Progesterone | Bipolar disorder | Women only | IVW | 0.24 | 1 | 0.624 |
| Schizophrenia | Testosterone [pre-menopause] | Women only | MR Egger | 20.91 | 16 | 0.182 |
| Schizophrenia | Testosterone [pre-menopause] | Women only | IVW | 29.27 | 17 | 0.032 |
| Schizophrenia | Testosterone [post-menopause] | Women only | MR Egger | 20.55 | 16 | 0.196 |
| Schizophrenia | Testosterone [post-menopause] | Women only | IVW | 20.60 | 17 | 0.245 |
| Schizophrenia | Testosterone [overall] | Women only | MR Egger | 65.44 | 20 | 9.9E-07 |
| Schizophrenia | Testosterone [overall] | Women only | IVW | 65.95 | 21 | 1.5E-06 |
| Schizophrenia | SHBG [pre-menopause] | Women only | MR Egger | 9.83 | 16 | 0.875 |
| Schizophrenia | SHBG [pre-menopause] | Women only | IVW | 10.18 | 17 | 0.896 |
| Schizophrenia | SHBG [post-menopause] | Women only | MR Egger | 29.96 | 16 | 0.018 |
| Schizophrenia | SHBG [post-menopause] | Women only | IVW | 30.35 | 17 | 0.024 |
| Schizophrenia | Progesterone | Women only | MR Egger | 1.22 | 20 | 0.999999999 |
| Schizophrenia | Progesterone | Women only | IVW | 1.50 | 21 | 0.999999998 |
| Schizophrenia | Oestrogen [pre-menopause] | Women only | MR Egger | 13.90 | 16 | 0.606 |
| Schizophrenia | Oestrogen [pre-menopause] | Women only | IVW | 15.07 | 17 | 0.591 |
| Schizophrenia | Oestrogen [post-menopause] | Women only | MR Egger | 18.64 | 16 | 0.288 |
| Schizophrenia | Oestrogen [post-menopause] | Women only | IVW | 19.25 | 17 | 0.314 |
| Schizophrenia | Oestrogen [overall] | Women only | MR Egger | 23.42 | 16 | 0.103 |
| Schizophrenia | Oestrogen [overall] | Women only | IVW | 26.52 | 17 | 0.065 |
| Schizophrenia | Age of menopause | Women only | MR Egger | 6.11 | 8 | 0.635 |
| Schizophrenia | Age of menopause | Women only | IVW | 8.55 | 9 | 0.480 |
| Schizophrenia | Age of menarche | Women only | MR Egger | 8.50 | 8 | 0.386 |
| Schizophrenia | Age of menarche | Women only | IVW | 12.61 | 9 | 0.181 |
| Schizophrenia | Testosterone | Men only | MR Egger | 294.74 | 51 | 5.2E-36 |
| Schizophrenia | Testosterone | Men only | IVW | 297.15 | 52 | 4.6E-36 |
| Schizophrenia | SHBG | Men only | MR Egger | 189.87 | 51 | 7.2E-18 |
| Schizophrenia | SHBG | Men only | IVW | 190.79 | 52 | 9.9E-18 |
| Schizophrenia | Oestrogen | Men only | MR Egger | 53.97 | 51 | 0.361 |
| Schizophrenia | Oestrogen | Men only | IVW | 54.76 | 52 | 0.370 |
| SHBG | Schizophrenia | Men only | MR Egger | 396.62 | 175 | 3.1E-19 |
| SHBG | Schizophrenia | Men only | IVW | 397.22 | 176 | 3.9E-19 |
| SHBG | Depressive disorder | Men only | MR Egger | 110.85 | 121 | 0.735 |
| SHBG | Depressive disorder | Men only | IVW | 115.22 | 122 | 0.655 |
| SHBG | Bipolar disorder | Men only | MR Egger | 173.19 | 123 | 0.002 |
| SHBG | Bipolar disorder | Men only | IVW | 173.19 | 124 | 0.002 |
| SHBG | Schizophrenia | Sexes combined | MR Egger | 907.83 | 304 | 2.0E-61 |
| SHBG | Schizophrenia | Sexes combined | IVW | 907.98 | 305 | 3.4E-61 |
| SHBG | Depressive disorder | Sexes combined | MR Egger | 420.36 | 333 | 8.1E-04 |
| SHBG | Depressive disorder | Sexes combined | IVW | 423.02 | 334 | 0.001 |
| SHBG | Bipolar disorder | Sexes combined | MR Egger | 736.28 | 305 | 1.2E-37 |
| SHBG | Bipolar disorder | Sexes combined | IVW | 736.96 | 306 | 1.5E-37 |
| SHBG [post-menopause] | Schizophrenia | Women only | MR Egger | 150.62 | 80 | 3.1E-06 |
| SHBG [post-menopause] | Schizophrenia | Women only | IVW | 151.95 | 81 | 3.1E-06 |
| SHBG [post-menopause] | Depressive disorder | Women only | MR Egger | 36.65 | 51 | 0.935 |
| SHBG [post-menopause] | Depressive disorder | Women only | IVW | 38.17 | 52 | 0.924 |
| SHBG [post-menopause] | Bipolar disorder | Women only | MR Egger | 68.95 | 52 | 0.058 |
| SHBG [post-menopause] | Bipolar disorder | Women only | IVW | 71.31 | 53 | 0.047 |
| SHBG [pre-menopause] | Schizophrenia | Women only | MR Egger | 35.41 | 20 | 0.018 |
| SHBG [pre-menopause] | Schizophrenia | Women only | IVW | 37.10 | 21 | 0.016 |
| SHBG [pre-menopause] | Depressive disorder | Women only | MR Egger | 7.97 | 14 | 0.891 |
| SHBG [pre-menopause] | Depressive disorder | Women only | IVW | 7.97 | 15 | 0.925 |
| SHBG [pre-menopause] | Bipolar disorder | Women only | MR Egger | 21.85 | 13 | 0.058 |
| SHBG [pre-menopause] | Bipolar disorder | Women only | IVW | 22.36 | 14 | 0.072 |
| Testosterone [overall] | Schizophrenia | Women only | MR Egger | 274.52 | 176 | 2.8E-06 |
| Testosterone [overall] | Schizophrenia | Women only | IVW | 274.55 | 177 | 3.6E-06 |
| Testosterone [overall] | Depressive disorder | Women only | MR Egger | 161.72 | 120 | 0.007 |
| Testosterone [overall] | Depressive disorder | Women only | IVW | 161.75 | 121 | 0.008 |
| Testosterone [overall] | Bipolar disorder | Women only | MR Egger | 146.81 | 126 | 0.099 |
| Testosterone [overall] | Bipolar disorder | Women only | IVW | 147.87 | 127 | 0.099 |
| Testosterone | Schizophrenia | Men only | MR Egger | 334.20 | 145 | 5.7E-17 |
| Testosterone | Schizophrenia | Men only | IVW | 334.49 | 146 | 8.0E-17 |
| Testosterone | Depressive disorder | Men only | MR Egger | 76.68 | 97 | 0.937 |
| Testosterone | Depressive disorder | Men only | IVW | 76.70 | 98 | 0.945 |
| Testosterone | Bipolar disorder | Men only | MR Egger | 112.04 | 98 | 0.157 |
| Testosterone | Bipolar disorder | Men only | IVW | 112.60 | 99 | 0.165 |
| Testosterone | Schizophrenia | Sexes combined | MR Egger | 463.52 | 157 | 5.1E-32 |
| Testosterone | Schizophrenia | Sexes combined | IVW | 464.30 | 158 | 6.8E-32 |
| Testosterone | Depressive disorder | Sexes combined | MR Egger | 196.34 | 164 | 4.3E-02 |
| Testosterone | Depressive disorder | Sexes combined | IVW | 196.42 | 165 | 0.048 |
| Testosterone | Bipolar disorder | Sexes combined | MR Egger | 320.08 | 157 | 3.2E-13 |
| Testosterone | Bipolar disorder | Sexes combined | IVW | 320.37 | 158 | 4.2E-13 |

#### Multivariable MR analyses

**Table S9.** Multivariable MR analysis for the effect of sex hormone traits on SMI risk (main + exploratory analyses)

| **Exposure** | **Mediator** | **Outcome** | **Analysis** | **Effect** | **beta** | **se** | **pval** |
| --- | --- | --- | --- | --- | --- | --- | --- |
| Testosterone | Cortisol | Schizophrenia | Men only | Effect of exposure on outcome | 0.07 | 0.05 | 0.148 |
| Testosterone | Cortisol | Schizophrenia | Men only | Effect of mediator on outcome | -0.15 | 0.11 | 0.163 |
| Testosterone | CRP | Schizophrenia | Men only | Effect of exposure on outcome | -0.01 | 0.09 | 0.932 |
| Testosterone | CRP | Schizophrenia | Men only | Effect of mediator on outcome | -0.13 | 0.04 | 0.003 |
| Testosterone | SI | Schizophrenia | Men only | Effect of exposure on outcome | 0.08 | 0.06 | 0.132 |
| Testosterone | SI | Schizophrenia | Men only | Effect of mediator on outcome | 0.56 | 0.19 | 0.004 |
| Testosterone | DrnkWk | Schizophrenia | Men only | Effect of exposure on outcome | 0.07 | 0.05 | 0.175 |
| Testosterone | DrnkWk | Schizophrenia | Men only | Effect of mediator on outcome | -0.02 | 0.23 | 0.921 |
| Testosterone | AD | Schizophrenia | Men only | Effect of exposure on outcome | 0.11 | 0.05 | 0.045 |
| Testosterone | AD | Schizophrenia | Men only | Effect of mediator on outcome | -0.03 | 0.16 | 0.837 |
| SHBG [pre-menopause] | Cortisol | Depressive disorder | Women only | Effect of exposure on outcome | 0.15 | 0.05 | 0.014 |
| SHBG [pre-menopause] | Cortisol | Depressive disorder | Women only | Effect of mediator on outcome | -0.05 | 0.10 | 0.657 |
| SHBG [pre-menopause] | CRP | Depressive disorder | Women only | Effect of exposure on outcome | 0.14 | 0.08 | 0.065 |
| SHBG [pre-menopause] | CRP | Depressive disorder | Women only | Effect of mediator on outcome | 0.02 | 0.04 | 0.569 |
| SHBG [pre-menopause] | SI | Depressive disorder | Women only | Effect of exposure on outcome | 0.13 | 0.07 | 0.066 |
| SHBG [pre-menopause] | SI | Depressive disorder | Women only | Effect of mediator on outcome | 0.19 | 0.22 | 0.399 |
| SHBG [pre-menopause] | DrnkWk | Depressive disorder | Women only | Effect of exposure on outcome | 0.19 | 0.06 | 0.007 |
| SHBG [pre-menopause] | DrnkWk | Depressive disorder | Women only | Effect of mediator on outcome | -0.91 | 0.34 | 0.015 |
| SHBG [pre-menopause] | AD | Depressive disorder | Women only | Effect of exposure on outcome | 0.16 | 0.06 | 0.022 |
| SHBG [pre-menopause] | AD | Depressive disorder | Women only | Effect of mediator on outcome | -0.36 | 0.42 | 0.400 |
| SHBG [post-menopause] | Cortisol | Depressive disorder | Women only | Effect of exposure on outcome | 0.21 | 0.05 | 1.6E-04 |
| SHBG [post-menopause] | Cortisol | Depressive disorder | Women only | Effect of mediator on outcome | 4.1E-03 | 0.11 | 0.971 |
| SHBG [post-menopause] | CRP | Depressive disorder | Women only | Effect of exposure on outcome | 0.20 | 0.07 | 0.004 |
| SHBG [post-menopause] | CRP | Depressive disorder | Women only | Effect of mediator on outcome | 0.02 | 0.04 | 0.566 |
| SHBG [post-menopause] | SI | Depressive disorder | Women only | Effect of exposure on outcome | 0.20 | 0.06 | 0.001 |
| SHBG [post-menopause] | SI | Depressive disorder | Women only | Effect of mediator on outcome | 0.16 | 0.21 | 0.459 |
| SHBG [post-menopause] | DrnkWk | Depressive disorder | Women only | Effect of exposure on outcome | 0.22 | 0.05 | 1.7E-04 |
| SHBG [post-menopause] | DrnkWk | Depressive disorder | Women only | Effect of mediator on outcome | -0.53 | 0.33 | 0.118 |
| SHBG [post-menopause] | AD | Depressive disorder | Women only | Effect of exposure on outcome | 0.22 | 0.05 | 1.5E-04 |
| SHBG [post-menopause] | AD | Depressive disorder | Women only | Effect of mediator on outcome | -0.18 | 0.28 | 0.530 |

*CRP = C-reactive protein, SI = smoking initiation, DrnkWk = alcohol drinks per week, AD = alcohol dependence*

**Table S10.** Multivariable MR analysis for the effect of liability to SMI on sex hormone traits (main + exploratory analyses)

| **Exposure** | **Mediator** | **Outcome** | **Analysis** | **Effect** | **beta** | **se** | **pval** |
| --- | --- | --- | --- | --- | --- | --- | --- |
| Schizophrenia | Cortisol | Testosterone | Sexes combined | Effect of exposure on outcome | 4.0E-03 | 2.8E-03 | 0.153 |
| Schizophrenia | Cortisol | Testosterone | Sexes combined | Effect of mediator on outcome | -1.8E-05 | 0.01 | 0.999 |
| Schizophrenia | CRP | Testosterone | Sexes combined | Effect of exposure on outcome | 0.01 | 0.01 | 0.324 |
| Schizophrenia | CRP | Testosterone | Sexes combined | Effect of mediator on outcome | -0.02 | 0.01 | 0.043 |
| Schizophrenia | BMI | Testosterone | Sexes combined | Effect of exposure on outcome | 0.01 | 4.2E-03 | 0.103 |
| Schizophrenia | BMI | Testosterone | Sexes combined | Effect of mediator on outcome | -0.03 | 0.01 | 0.003 |
| Schizophrenia | WHR | Testosterone | Sexes combined | Effect of exposure on outcome | 0.01 | 4.4E-03 | 0.068 |
| Schizophrenia | WHR | Testosterone | Sexes combined | Effect of mediator on outcome | -0.05 | 0.02 | 0.012 |
| Schizophrenia | SI | Testosterone | Sexes combined | Effect of exposure on outcome | 0.01 | 2.9E-03 | 0.078 |
| Schizophrenia | SI | Testosterone | Sexes combined | Effect of mediator on outcome | -0.03 | 0.02 | 0.135 |
| Schizophrenia | DrnkWk | Testosterone | Sexes combined | Effect of exposure on outcome | 4.7E-03 | 3.9E-03 | 0.234 |
| Schizophrenia | DrnkWk | Testosterone | Sexes combined | Effect of mediator on outcome | 0.08 | 0.04 | 0.040 |
| Schizophrenia | Cortisol | SHBG [post-menopause] | Women only | Effect of exposure on outcome | 0.05 | 0.02 | 0.009 |
| Schizophrenia | Cortisol | SHBG [post-menopause] | Women only | Effect of mediator on outcome | -0.08 | 0.06 | 0.199 |
| Schizophrenia | CRP | SHBG [post-menopause] | Women only | Effect of exposure on outcome | 0.04 | 0.05 | 0.424 |
| Schizophrenia | CRP | SHBG [post-menopause] | Women only | Effect of mediator on outcome | -0.07 | 0.05 | 0.115 |
| Schizophrenia | BMI | SHBG [post-menopause] | Women only | Effect of exposure on outcome | 0.05 | 0.03 | 0.058 |
| Schizophrenia | BMI | SHBG [post-menopause] | Women only | Effect of mediator on outcome | -0.15 | 0.04 | 4.6E-04 |
| Schizophrenia | WHR | SHBG [post-menopause] | Women only | Effect of exposure on outcome | 0.06 | 0.03 | 0.033 |
| Schizophrenia | WHR | SHBG [post-menopause] | Women only | Effect of mediator on outcome | -0.44 | 0.05 | 8.1E-10 |
| Schizophrenia | SI | SHBG [post-menopause] | Women only | Effect of exposure on outcome | 0.05 | 0.02 | 0.002 |
| Schizophrenia | SI | SHBG [post-menopause] | Women only | Effect of mediator on outcome | 0.08 | 0.07 | 0.228 |
| Schizophrenia | DrnkWk | SHBG [post-menopause] | Women only | Effect of exposure on outcome | 0.04 | 0.04 | 0.266 |
| Schizophrenia | DrnkWk | SHBG [post-menopause] | Women only | Effect of mediator on outcome | 0.38 | 0.27 | 0.175 |
| Bipolar disorder | Cortisol | SHBG | Sexes combined | Effect of exposure on outcome | 0.01 | 0.01 | 0.053 |
| Bipolar disorder | Cortisol | SHBG | Sexes combined | Effect of mediator on outcome | -0.07 | 0.02 | 0.005 |
| Bipolar disorder | CRP | SHBG | Sexes combined | Effect of exposure on outcome | -0.01 | 0.03 | 0.639 |
| Bipolar disorder | CRP | SHBG | Sexes combined | Effect of mediator on outcome | -0.03 | 0.02 | 0.114 |
| Bipolar disorder | BMI | SHBG | Sexes combined | Effect of exposure on outcome | 0.01 | 0.01 | 0.179 |
| Bipolar disorder | BMI | SHBG | Sexes combined | Effect of mediator on outcome | -0.08 | 0.01 | 4.4E-09 |
| Bipolar disorder | WHR | SHBG | Sexes combined | Effect of exposure on outcome | 0.01 | 0.02 | 0.598 |
| Bipolar disorder | WHR | SHBG | Sexes combined | Effect of mediator on outcome | -0.10 | 0.03 | 0.002 |
| Bipolar disorder | SI | SHBG | Sexes combined | Effect of exposure on outcome | 0.01 | 0.01 | 0.129 |
| Bipolar disorder | SI | SHBG | Sexes combined | Effect of mediator on outcome | 0.01 | 0.02 | 0.684 |
| Bipolar disorder | DrnkWk | SHBG | Sexes combined | Effect of exposure on outcome | 2.4E-03 | 0.01 | 0.856 |
| Bipolar disorder | DrnkWk | SHBG | Sexes combined | Effect of mediator on outcome | 0.09 | 0.08 | 0.218 |

*CRP = C-reactive protein, SI = smoking initiation, DrnkWk = alcohol drinks per week, AD = alcohol dependence, WHR = waist-hip-ratio*

**Table S11.** Conditional F-statistic across all SNPs included in the genetic instruments used in the multivariable MR analysis for the effect of sex hormone traits to SMI risk (main + exploratory analyses)

| **Exposure** | **Mediator** | **Outcome** | **Analysis** | **F Exposure** | **F Mediator** |
| --- | --- | --- | --- | --- | --- |
| Testosterone | Cortisol | Schizophrenia | Men only | 16.03 | 2.54 |
| Testosterone | CRP | Schizophrenia | Men only | 45.53 | 116.88 |
| Testosterone | SI | Schizophrenia | Men only | 85.94 | 12.75 |
| Testosterone | DrnkWk | Schizophrenia | Men only | 105.61 | 5.25 |
| Testosterone | AD | Schizophrenia | Men only | 72.75 | 1.92 |
| SHBG [pre-menopause] | Cortisol | Depressive disorder | Women only | 55.98 | 10.31 |
| SHBG [pre-menopause] | CRP | Depressive disorder | Women only | 15.45 | 131.88 |
| SHBG [pre-menopause] | SI | Depressive disorder | Women only | 28.63 | 29.83 |
| SHBG [pre-menopause] | DrnkWk | Depressive disorder | Women only | 41.02 | 9.81 |
| SHBG [pre-menopause] | AD | Depressive disorder | Women only | 6.07 | 2.68 |
| SHBG [post-menopause] | Cortisol | Depressive disorder | Women only | 60.01 | 4.10 |
| SHBG [post-menopause] | CRP | Depressive disorder | Women only | 33.35 | 131.93 |
| SHBG [post-menopause] | SI | Depressive disorder | Women only | 46.94 | 17.35 |
| SHBG [post-menopause] | DrnkWk | Depressive disorder | Women only | 65.39 | 4.75 |
| SHBG [post-menopause] | AD | Depressive disorder | Women only | 7.58 | 1.73 |

*CRP = C-reactive protein, SI = smoking initiation, DrnkWk = alcohol drinks per week, AD = alcohol dependence*

**Table S12.** Conditional F-statistic across all SNPs included in the genetic instruments used in the multivariable MR analysis for the effect of liability to SMI on sex hormone traits

| **Exposure** | **Outcome** | **Mediator** | **Analysis** | **F Exposure** | **F Mediator** |
| --- | --- | --- | --- | --- | --- |
| Schizophrenia | T | Cortisol | Sexes combined | 35.88 | 2.05 |
| Schizophrenia | T | CRP | Sexes combined | 31.37 | 71.28 |
| Schizophrenia | T | BMI | Sexes combined | 26.50 | 32.05 |
| Schizophrenia | T | WHR | Sexes combined | 33.86 | 13.34 |
| Schizophrenia | T | SI | Sexes combined | 34.33 | 12.52 |
| Schizophrenia | T | DrnkWk | Sexes combined | 42.14 | 5.15 |
| Schizophrenia | SHBG [post-menopause] | Cortisol | Women only | 23.64 | 8.63 |
| Schizophrenia | SHBG [post-menopause] | CRP | Women only | 8.61 | 161.84 |
| Schizophrenia | SHBG [post-menopause] | BMI | Women only | 6.95 | 49.40 |
| Schizophrenia | SHBG [post-menopause] | WHR | Women only | 12.21 | 37.50 |
| Schizophrenia | SHBG [post-menopause] | SI | Women only | 13.89 | 31.60 |
| Schizophrenia | SHBG [post-menopause] | DrnkWk | Women only | 25.24 | 13.43 |
| Bipolar disorder | SHBG | Cortisol | Sexes combined | 36.56 | 4.27 |
| Bipolar disorder | SHBG | CRP | Sexes combined | 13.36 | 140.57 |
| Bipolar disorder | SHBG | BMI | Sexes combined | 8.53 | 49.29 |
| Bipolar disorder | SHBG | WHR | Sexes combined | 14.68 | 28.11 |
| Bipolar disorder | SHBG | SI | Sexes combined | 22.30 | 24.06 |
| Bipolar disorder | SHBG | DrnkWk | Sexes combined | 32.69 | 11.84 |

*CRP = C-reactive protein, SI = smoking initiation, DrnkWk = alcohol drinks per week, AD = alcohol dependence, WHR = waist-hip-ratio*

**Table S13.** Cochran’s Q-statistic for the effect of sex hormone traits on SMI risk, indicating evidence for heterogeneity

| **Exposure** | **Analysis** | **Mediator** | **Outcome** | **Q statistic** | **Q pval** |
| --- | --- | --- | --- | --- | --- |
| Testosterone | Men only | Cortisol | Schizophrenia | 322.04 | 1.8E-15 |
| Testosterone | Men only | CRP | Schizophrenia | 203.51 | 4.7E-10 |
| Testosterone | Men only | SI | Schizophrenia | 508.83 | 5.0E-32 |
| Testosterone | Men only | DrnkWk | Schizophrenia | 376.89 | 4.2E-21 |
| Testosterone | Men only | AD | Schizophrenia | 313.65 | 3.7E-15 |
| SHBG [pre-menopause] | Women only | Cortisol | Depressive disorder | 8.34 | 0.938 |
| SHBG [pre-menopause] | Women only | CRP | Depressive disorder | 27.34 | 0.950 |
| SHBG [pre-menopause] | Women only | SI | Depressive disorder | 35.95 | 0.610 |
| SHBG [pre-menopause] | Women only | DrnkWk | Depressive disorder | 12.54 | 0.818 |
| SHBG [pre-menopause] | Women only | AD | Depressive disorder | 8.26 | 0.826 |
| SHBG [post-menopause] | Women only | Cortisol | Depressive disorder | 38.62 | 0.931 |
| SHBG [post-menopause] | Women only | CRP | Depressive disorder | 38.27 | 0.979 |
| SHBG [post-menopause] | Women only | SI | Depressive disorder | 66.24 | 0.780 |
| SHBG [post-menopause] | Women only | DrnkWk | Depressive disorder | 45.36 | 0.820 |
| SHBG [post-menopause] | Women only | AD | Depressive disorder | 36.51 | 0.906 |

*CRP = C-reactive protein, SI = smoking initiation, DrnkWk = alcohol drinks per week, AD = alcohol dependence*

**Table S14.** Cochran’s Q-statistic for the effect of liability to SMI on sex hormone traits, indicating evidence for heterogeneity

| **Exposure** | **Analysis** | **Mediator** | **Outcome** | **Q statistic** | **Q pval** |
| --- | --- | --- | --- | --- | --- |
| Schizophrenia | Sexes combined | Cortisol | Testosterone | 461.06 | 7.8E-28 |
| Schizophrenia | Sexes combined | CRP | Testosterone | 986.01 | 2.6E-133 |
| Schizophrenia | Sexes combined | BMI | Testosterone | 655.89 | 2.5E-57 |
| Schizophrenia | Sexes combined | WHR | Testosterone | 461.75 | 5.6E-41 |
| Schizophrenia | Sexes combined | SI | Testosterone | 646.30 | 1.0E-44 |
| Schizophrenia | Sexes combined | DrnkWk | Testosterone | 979.18 | 2.4E-109 |
| Schizophrenia | Women only | Cortisol | SHBG [post-menopause] | 31.84 | 0.023 |
| Schizophrenia | Women only | CRP | SHBG [post-menopause] | 581.30 | 1.1E-92 |
| Schizophrenia | Women only | BMI | SHBG [post-menopause] | 313.67 | 8.5E-27 |
| Schizophrenia | Women only | WHR | SHBG [post-menopause] | 74.92 | 1.0E-05 |
| Schizophrenia | Women only | SI | SHBG [post-menopause] | 101.44 | 1.4E-04 |
| Schizophrenia | Women only | DrnkWk | SHBG [post-menopause] | 230.73 | 1.1E-35 |
| Bipolar disorder | Sexes combined | Cortisol | SHBG | 293.12 | 6.8E-37 |
| Bipolar disorder | Sexes combined | CRP | SHBG | 3230.78 | 0.00 |
| Bipolar disorder | Sexes combined | BMI | SHBG | 883.44 | 3.3E-125 |
| Bipolar disorder | Sexes combined | WHR | SHBG | 816.33 | 4.3E-139 |
| Bipolar disorder | Sexes combined | SI | SHBG | 560.29 | 7.3E-70 |
| Bipolar disorder | Sexes combined | DrnkWk | SHBG | 1663.85 | 0.00 |

*CRP = C-reactive protein, SI = smoking initiation, DrnkWk = alcohol drinks per week, AD = alcohol dependence, WHR = waist-hip-ratio*

**Table S15.** MR-Egger intercepts for the effect of sex hormone traits on SMI risk, indicating evidence for horizontal pleiotropy

|  |  |  |  |  | **No orientation** | | | **Orientation X1** | | | **Orientation X2** | | | |
| --- | --- | --- | --- | --- | --- | --- | --- | --- | --- | --- | --- | --- | --- | --- |
| **Exposure** | **Mediator** | **Outcome** | **Analysis** | **Effect** | **beta** | **se** | **pval** | **beta** | **se** | **pval** | | **beta** | **se** | **pval** |
| Testosterone | Cortisol | Schizophrenia | Men only | Intercept | 2.0E-03 | 3.0E-03 | 0.493 | 1.6E-03 | 2.9E-03 | 0.596 | | 5.9E-06 | 2.5E-03 | 0.998 |
| Testosterone | Cortisol | Schizophrenia | Men only | Effect of exposure on outcome | 0.03 | 0.09 | 0.753 | 0.04 | 0.08 | 0.645 | | 0.07 | 0.05 | 0.150 |
| Testosterone | Cortisol | Schizophrenia | Men only | Effect of mediator on outcome | -0.17 | 0.11 | 0.128 | -0.16 | 0.11 | 0.160 | | -0.15 | 0.15 | 0.312 |
| Testosterone | CRP | Schizophrenia | Men only | Intercept | -6.7E-04 | 2.6E-03 | 0.796 | 1.1E-03 | 3.7E-03 | 0.766 | | 2.6E-03 | 2.6E-03 | 0.335 |
| Testosterone | CRP | Schizophrenia | Men only | Effect of exposure on outcome | 0.01 | 0.11 | 0.926 | -0.05 | 0.16 | 0.769 | | 0.02 | 0.09 | 0.867 |
| Testosterone | CRP | Schizophrenia | Men only | Effect of mediator on outcome | -0.13 | 0.04 | 0.003 | -0.13 | 0.04 | 0.004 | | -0.16 | 0.06 | 0.004 |
| Testosterone | SI | Schizophrenia | Men only | Intercept | 1.8E-03 | 2.3E-03 | 0.442 | 3.0E-03 | 2.5E-03 | 0.223 | | -1.7E-03 | 2.5E-03 | 0.499 |
| Testosterone | SI | Schizophrenia | Men only | Effect of exposure on outcome | 0.04 | 0.08 | 0.559 | 0.01 | 0.08 | 0.869 | | 0.08 | 0.06 | 0.136 |
| Testosterone | SI | Schizophrenia | Men only | Effect of mediator on outcome | 0.56 | 0.19 | 0.003 | 0.55 | 0.19 | 0.004 | | 0.69 | 0.27 | 0.012 |
| Testosterone | DrnkWk | Schizophrenia | Men only | Intercept | 3.2E-03 | 2.9E-03 | 0.258 | 2.0E-03 | 3.0E-03 | 0.500 | | 2.4E-03 | 2.1E-03 | 0.259 |
| Testosterone | DrnkWk | Schizophrenia | Men only | Effect of exposure on outcome | 1.0E-03 | 0.08 | 0.990 | 0.03 | 0.08 | 0.757 | | 0.07 | 0.05 | 0.193 |
| Testosterone | DrnkWk | Schizophrenia | Men only | Effect of mediator on outcome | -0.05 | 0.23 | 0.825 | -0.03 | 0.23 | 0.894 | | -0.18 | 0.27 | 0.508 |
| Testosterone | AD | Schizophrenia | Men only | Intercept | -1.1E-03 | 3.1E-03 | 0.726 | -9.4E-04 | 3.1E-03 | 0.763 | | 4.9E-05 | 2.3E-03 | 0.983 |
| Testosterone | AD | Schizophrenia | Men only | Effect of exposure on outcome | 0.13 | 0.09 | 0.149 | 0.13 | 0.09 | 0.162 | | 0.11 | 0.05 | 0.046 |
| Testosterone | AD | Schizophrenia | Men only | Effect of mediator on outcome | -0.03 | 0.16 | 0.835 | -0.04 | 0.16 | 0.821 | | -0.04 | 0.21 | 0.864 |
| SHBG [pre-menopause] | Cortisol | Depressive disorder | Women only | Intercept | 1.2E-03 | 4.3E-03 | 0.778 | 4.7E-04 | 0.01 | 0.961 | | 1.9E-03 | 0.01 | 0.726 |
| SHBG [pre-menopause] | Cortisol | Depressive disorder | Women only | Effect of exposure on outcome | 0.14 | 0.06 | 0.027 | 0.14 | 0.13 | 0.275 | | 0.15 | 0.06 | 0.018 |
| SHBG [pre-menopause] | Cortisol | Depressive disorder | Women only | Effect of mediator on outcome | -0.06 | 0.12 | 0.609 | -0.04 | 0.14 | 0.774 | | -0.08 | 0.14 | 0.579 |
| SHBG [pre-menopause] | CRP | Depressive disorder | Women only | Intercept | -3.0E-03 | 2.3E-03 | 0.208 | -2.7E-04 | 3.3E-03 | 0.934 | | -2.3E-03 | 3.5E-03 | 0.518 |
| SHBG [pre-menopause] | CRP | Depressive disorder | Women only | Effect of exposure on outcome | 0.15 | 0.08 | 0.056 | 0.15 | 0.11 | 0.162 | | 0.14 | 0.08 | 0.072 |
| SHBG [pre-menopause] | CRP | Depressive disorder | Women only | Effect of mediator on outcome | 0.02 | 0.04 | 0.612 | 0.02 | 0.04 | 0.573 | | 0.05 | 0.06 | 0.392 |
| SHBG [pre-menopause] | SI | Depressive disorder | Women only | Intercept | 4.9E-03 | 3.1E-03 | 0.120 | -1.5E-03 | 4.0E-03 | 0.708 | | -0.01 | 0.01 | 0.382 |
| SHBG [pre-menopause] | SI | Depressive disorder | Women only | Effect of exposure on outcome | 0.12 | 0.07 | 0.111 | 0.16 | 0.09 | 0.098 | | 0.13 | 0.07 | 0.069 |
| SHBG [pre-menopause] | SI | Depressive disorder | Women only | Effect of mediator on outcome | 0.27 | 0.23 | 0.245 | 0.16 | 0.24 | 0.496 | | 0.54 | 0.45 | 0.241 |
| SHBG [pre-menopause] | DrnkWk | Depressive disorder | Women only | Intercept | 2.4E-03 | 4.2E-03 | 0.571 | -0.01 | 0.01 | 0.315 | | -5.2E-04 | 0.01 | 0.931 |
| SHBG [pre-menopause] | DrnkWk | Depressive disorder | Women only | Effect of exposure on outcome | 0.18 | 0.07 | 0.015 | 0.28 | 0.11 | 0.018 | | 0.19 | 0.06 | 0.009 |
| SHBG [pre-menopause] | DrnkWk | Depressive disorder | Women only | Effect of mediator on outcome | -0.90 | 0.35 | 0.018 | -0.79 | 0.36 | 0.042 | | -0.88 | 0.51 | 0.104 |
| SHBG [pre-menopause] | AD | Depressive disorder | Women only | Intercept | -3.0E-04 | 4.9E-03 | 0.952 | -5.8E-04 | 0.01 | 0.962 | | -0.01 | 0.01 | 0.383 |
| SHBG [pre-menopause] | AD | Depressive disorder | Women only | Effect of exposure on outcome | 0.16 | 0.07 | 0.030 | 0.17 | 0.15 | 0.295 | | 0.16 | 0.06 | 0.028 |
| SHBG [pre-menopause] | AD | Depressive disorder | Women only | Effect of mediator on outcome | -0.36 | 0.44 | 0.437 | -0.35 | 0.47 | 0.460 | | 0.04 | 0.61 | 0.955 |
| SHBG [post-menopause] | Cortisol | Depressive disorder | Women only | Intercept | 3.3E-03 | 2.6E-03 | 0.204 | 0.01 | 0.01 | 0.122 | | 4.5E-03 | 3.0E-03 | 0.146 |
| SHBG [post-menopause] | Cortisol | Depressive disorder | Women only | Effect of exposure on outcome | 0.20 | 0.05 | 0.001 | 0.06 | 0.11 | 0.607 | | 0.21 | 0.05 | 1.3E-04 |
| SHBG [post-menopause] | Cortisol | Depressive disorder | Women only | Effect of mediator on outcome | -0.04 | 0.12 | 0.748 | 0.06 | 0.12 | 0.610 | | -0.13 | 0.14 | 0.383 |
| SHBG [post-menopause] | CRP | Depressive disorder | Women only | Intercept | -8.2E-04 | 2.0E-03 | 0.687 | 4.7E-04 | 3.2E-03 | 0.883 | | -1.9E-03 | 2.8E-03 | 0.507 |
| SHBG [post-menopause] | CRP | Depressive disorder | Women only | Effect of exposure on outcome | 0.20 | 0.07 | 0.004 | 0.19 | 0.10 | 0.075 | | 0.19 | 0.07 | 6.2E-03 |
| SHBG [post-menopause] | CRP | Depressive disorder | Women only | Effect of mediator on outcome | 0.02 | 0.04 | 0.584 | 0.02 | 0.04 | 0.564 | | 0.05 | 0.05 | 0.386 |
| SHBG [post-menopause] | SI | Depressive disorder | Women only | Intercept | 4.5E-03 | 2.2E-03 | 0.045 | 4.1E-04 | 3.4E-03 | 0.904 | | -0.01 | 3.4E-03 | 0.052 |
| SHBG [post-menopause] | SI | Depressive disorder | Women only | Effect of exposure on outcome | 0.18 | 0.06 | 0.002 | 0.20 | 0.09 | 0.030 | | 0.19 | 0.06 | 0.001 |
| SHBG [post-menopause] | SI | Depressive disorder | Women only | Effect of mediator on outcome | 0.24 | 0.21 | 0.258 | 0.15 | 0.21 | 0.467 | | 0.65 | 0.32 | 0.048 |
| SHBG [post-menopause] | DrnkWk | Depressive disorder | Women only | Intercept | 3.7E-03 | 2.6E-03 | 0.159 | 2.4E-03 | 0.01 | 0.647 | | 0.01 | 3.1E-03 | 0.094 |
| SHBG [post-menopause] | DrnkWk | Depressive disorder | Women only | Effect of exposure on outcome | 0.20 | 0.06 | 0.001 | 0.18 | 0.11 | 0.117 | | 0.21 | 0.05 | 2.9E-04 |
| SHBG [post-menopause] | DrnkWk | Depressive disorder | Women only | Effect of mediator on outcome | -0.52 | 0.33 | 0.118 | -0.55 | 0.34 | 0.109 | | -0.93 | 0.40 | 0.025 |
| SHBG [post-menopause] | AD | Depressive disorder | Women only | Intercept | 3.6E-03 | 2.6E-03 | 0.171 | 0.01 | 0.01 | 0.181 | | -3.7E-04 | 3.8E-03 | 0.923 |
| SHBG [post-menopause] | AD | Depressive disorder | Women only | Effect of exposure on outcome | 0.21 | 0.06 | 3.4E-04 | 0.08 | 0.12 | 0.501 | | 0.22 | 0.06 | 1.9E-04 |
| SHBG [post-menopause] | AD | Depressive disorder | Women only | Effect of mediator on outcome | -0.26 | 0.28 | 0.368 | -0.20 | 0.28 | 0.476 | | -0.15 | 0.41 | 0.722 |

*CRP = C-reactive protein, SI = smoking initiation, DrnkWk = alcohol drinks per week, AD = alcohol dependence*

**Table S16.** MR-Egger intercepts for the effect of liability to SMI on sex hormone traits, indicating evidence for horizontal pleiotropy

|  |  |  |  |  | **No orientation** | | | **Orientation X1** | | | **Orientation X2** | | |
| --- | --- | --- | --- | --- | --- | --- | --- | --- | --- | --- | --- | --- | --- |
| **Exposure** | **Mediator** | **Outcome** | **Analysis** |  | **beta** | **se** | **pval** | **beta** | **se** | **pval** | **beta** | **se** | **pval** |
| Schizophrenia | Cortisol | Testosterone | Sexes combined | Intercept | -1.5E-04 | 1.8E-04 | 0.408 | 2.4E-04 | 6.4E-04 | 0.702 | 4.0E-04 | 2.5E-04 | 0.114 |
| Schizophrenia | Cortisol | Testosterone | Sexes combined | Effect of exposure on outcome | 4.0E-03 | 2.8E-03 | 0.152 | 3.6E-04 | 0.01 | 0.971 | 4.5E-03 | 2.8E-03 | 0.107 |
| Schizophrenia | Cortisol | Testosterone | Sexes combined | Effect of mediator on outcome | 9.4E-04 | 0.01 | 0.942 | 3.0E-04 | 0.01 | 0.981 | -0.02 | 0.02 | 0.269 |
| Schizophrenia | CRP | Testosterone | Sexes combined | Intercept | 2.5E-04 | 3.6E-04 | 0.489 | -2.3E-03 | 6.7E-04 | 0.001 | -2.0E-05 | 4.1E-04 | 0.961 |
| Schizophrenia | CRP | Testosterone | Sexes combined | Effect of exposure on outcome | 0.01 | 0.01 | 0.326 | 0.04 | 0.01 | 0.001 | 0.01 | 0.01 | 0.326 |
| Schizophrenia | CRP | Testosterone | Sexes combined | Effect of mediator on outcome | -0.02 | 0.01 | 0.049 | -0.03 | 0.01 | 0.004 | -0.02 | 0.01 | 0.084 |
| Schizophrenia | BMI | Testosterone | Sexes combined | Intercept | 2.2E-04 | 2.6E-04 | 0.392 | -8.5E-05 | 3.7E-04 | 0.817 | 1.3E-04 | 3.2E-04 | 0.694 |
| Schizophrenia | BMI | Testosterone | Sexes combined | Effect of exposure on outcome | 0.01 | 4.2E-03 | 0.103 | 0.01 | 0.01 | 0.254 | 0.01 | 4.2E-03 | 0.105 |
| Schizophrenia | BMI | Testosterone | Sexes combined | Effect of mediator on outcome | -0.04 | 0.01 | 0.004 | -0.03 | 0.01 | 0.004 | -0.04 | 0.02 | 0.024 |
| Schizophrenia | WHR | Testosterone | Sexes combined | Intercept | 1.7E-04 | 2.8E-04 | 0.546 | -1.9E-04 | 5.1E-04 | 0.716 | -5.8E-05 | 3.7E-04 | 0.874 |
| Schizophrenia | WHR | Testosterone | Sexes combined | Effect of exposure on outcome | 0.01 | 4.5E-03 | 0.069 | 0.01 | 0.01 | 0.223 | 0.01 | 4.5E-03 | 0.092 |
| Schizophrenia | WHR | Testosterone | Sexes combined | Effect of mediator on outcome | -0.05 | 0.02 | 0.011 | -0.05 | 0.02 | 0.011 | -0.04 | 0.03 | 0.100 |
| Schizophrenia | SI | Testosterone | Sexes combined | Intercept | -7.9E-05 | 1.7E-04 | 0.646 | -6.0E-05 | 4.3E-04 | 0.890 | -2.1E-04 | 2.5E-04 | 0.406 |
| Schizophrenia | SI | Testosterone | Sexes combined | Effect of exposure on outcome | 0.01 | 2.9E-03 | 0.078 | 0.01 | 0.01 | 0.400 | 0.01 | 2.9E-03 | 0.086 |
| Schizophrenia | SI | Testosterone | Sexes combined | Effect of mediator on outcome | -0.03 | 0.02 | 0.136 | -0.03 | 0.02 | 0.146 | -0.01 | 0.03 | 0.691 |
| Schizophrenia | DrnkWk | Testosterone | Sexes combined | Intercept | 4.8E-05 | 2.5E-04 | 0.849 | -1.9E-03 | 8.2E-04 | 0.024 | -4.0E-04 | 3.2E-04 | 0.216 |
| Schizophrenia | DrnkWk | Testosterone | Sexes combined | Effect of exposure on outcome | 4.7E-03 | 3.9E-03 | 0.236 | 0.03 | 0.01 | 0.012 | 0.01 | 0.00 | 0.183 |
| Schizophrenia | DrnkWk | Testosterone | Sexes combined | Effect of mediator on outcome | 0.07 | 0.04 | 0.042 | 0.09 | 0.04 | 0.018 | 0.11 | 0.05 | 0.017 |
| Schizophrenia | Cortisol | SHBG [post-menopause] | Women only | Intercept | 1.3E-03 | 1.6E-03 | 0.451 | 0.01 | 0.01 | 0.212 | 3.0E-03 | 1.9E-03 | 0.139 |
| Schizophrenia | Cortisol | SHBG [post-menopause] | Women only | Effect of exposure on outcome | 0.04 | 0.02 | 0.030 | -0.03 | 0.06 | 0.679 | 0.06 | 0.02 | 0.004 |
| Schizophrenia | Cortisol | SHBG [post-menopause] | Women only | Effect of mediator on outcome | -0.09 | 0.06 | 0.166 | 0.01 | 0.09 | 0.876 | -0.14 | 0.07 | 0.058 |
| Schizophrenia | CRP | SHBG [post-menopause] | Women only | Intercept | -7.6E-06 | 2.6E-03 | 0.998 | -3.3E-03 | 3.3E-03 | 0.330 | -0.01 | 3.3E-03 | 0.073 |
| Schizophrenia | CRP | SHBG [post-menopause] | Women only | Effect of exposure on outcome | 0.04 | 0.06 | 0.441 | 0.09 | 0.07 | 0.219 | 0.06 | 0.05 | 0.282 |
| Schizophrenia | CRP | SHBG [post-menopause] | Women only | Effect of mediator on outcome | -0.07 | 0.05 | 0.120 | -0.08 | 0.05 | 0.086 | 4.7E-04 | 0.06 | 0.994 |
| Schizophrenia | BMI | SHBG [post-menopause] | Women only | Intercept | -1.3E-03 | 2.3E-03 | 0.564 | -6.6E-04 | 1.4E-03 | 0.643 | -2.7E-03 | 2.6E-03 | 0.290 |
| Schizophrenia | BMI | SHBG [post-menopause] | Women only | Effect of exposure on outcome | 0.06 | 0.03 | 0.049 | 0.06 | 0.04 | 0.086 | 0.04 | 0.03 | 0.130 |
| Schizophrenia | BMI | SHBG [post-menopause] | Women only | Effect of mediator on outcome | -0.10 | 0.09 | 0.244 | -0.15 | 0.04 | 0.001 | -0.05 | 0.10 | 0.596 |
| Schizophrenia | WHR | SHBG [post-menopause] | Women only | Intercept | -2.3E-03 | 2.1E-03 | 0.281 | -3.6E-03 | 2.0E-03 | 0.087 | -4.8E-04 | 3.2E-03 | 0.881 |
| Schizophrenia | WHR | SHBG [post-menopause] | Women only | Effect of exposure on outcome | 0.08 | 0.03 | 0.018 | 0.11 | 0.04 | 0.007 | 0.06 | 0.03 | 0.051 |
| Schizophrenia | WHR | SHBG [post-menopause] | Women only | Effect of mediator on outcome | -0.39 | 0.07 | 1.5E-06 | -0.43 | 0.05 | 6.6E-10 | -0.42 | 0.11 | 3.8E-04 |
| Schizophrenia | SI | SHBG [post-menopause] | Women only | Intercept | 2.3E-03 | 9.0E-04 | 0.014 | 9.0E-04 | 1.5E-03 | 0.540 | -1.6E-03 | 2.1E-03 | 0.448 |
| Schizophrenia | SI | SHBG [post-menopause] | Women only | Effect of exposure on outcome | 0.04 | 0.02 | 0.009 | 0.04 | 0.03 | 0.119 | 0.05 | 0.02 | 0.002 |
| Schizophrenia | SI | SHBG [post-menopause] | Women only | Effect of mediator on outcome | 0.09 | 0.06 | 0.173 | 0.08 | 0.07 | 0.240 | 0.18 | 0.15 | 0.222 |
| Schizophrenia | DrnkWk | SHBG [post-menopause] | Women only | Intercept | 2.1E-03 | 3.5E-03 | 0.555 | -0.01 | 0.01 | 0.123 | 1.7E-03 | 4.6E-03 | 0.715 |
| Schizophrenia | DrnkWk | SHBG [post-menopause] | Women only | Effect of exposure on outcome | 0.04 | 0.04 | 0.416 | 0.16 | 0.08 | 0.062 | 0.05 | 0.04 | 0.260 |
| Schizophrenia | DrnkWk | SHBG [post-menopause] | Women only | Effect of mediator on outcome | 0.34 | 0.28 | 0.239 | 0.15 | 0.30 | 0.620 | 0.27 | 0.40 | 0.498 |
| Bipolar disorder | Cortisol | SHBG | Sexes combined | Intercept | -8.2E-05 | 4.4E-04 | 0.851 | 2.8E-03 | 1.7E-03 | 0.104 | 8.0E-04 | 5.2E-04 | 0.133 |
| Bipolar disorder | Cortisol | SHBG | Sexes combined | Effect of exposure on outcome | 0.01 | 0.01 | 0.060 | -0.03 | 0.03 | 0.278 | 0.01 | 0.01 | 0.071 |
| Bipolar disorder | Cortisol | SHBG | Sexes combined | Effect of mediator on outcome | -0.07 | 0.02 | 0.005 | -0.04 | 0.03 | 0.115 | -0.09 | 0.03 | 0.002 |
| Bipolar disorder | CRP | SHBG | Sexes combined | Intercept | -3.2E-04 | 9.9E-04 | 0.748 | 4.1E-04 | 1.4E-03 | 0.778 | -1.6E-03 | 1.3E-03 | 0.207 |
| Bipolar disorder | CRP | SHBG | Sexes combined | Effect of exposure on outcome | -0.01 | 0.03 | 0.624 | -0.02 | 0.04 | 0.600 | -0.01 | 0.03 | 0.670 |
| Bipolar disorder | CRP | SHBG | Sexes combined | Effect of mediator on outcome | -0.03 | 0.02 | 0.112 | -0.03 | 0.02 | 0.124 | -0.01 | 0.02 | 0.655 |
| Bipolar disorder | BMI | SHBG | Sexes combined | Intercept | 2.5E-04 | 5.9E-04 | 0.672 | 9.1E-04 | 3.7E-04 | 0.015 | 5.3E-05 | 6.2E-04 | 0.932 |
| Bipolar disorder | BMI | SHBG | Sexes combined | Effect of exposure on outcome | 0.01 | 0.01 | 0.166 | -5.0E-03 | 0.01 | 0.669 | 0.01 | 0.01 | 0.185 |
| Bipolar disorder | BMI | SHBG | Sexes combined | Effect of mediator on outcome | -0.09 | 0.02 | 2.0E-04 | -0.09 | 0.01 | 1.4E-13 | -0.08 | 0.02 | 0.001 |
| Bipolar disorder | WHR | SHBG | Sexes combined | Intercept | 2.2E-04 | 8.2E-04 | 0.792 | 4.1E-04 | 1.0E-03 | 0.683 | 2.1E-04 | 1.1E-03 | 0.853 |
| Bipolar disorder | WHR | SHBG | Sexes combined | Effect of exposure on outcome | 0.01 | 0.02 | 0.580 | 7.4E-04 | 0.02 | 0.975 | 0.01 | 0.02 | 0.615 |
| Bipolar disorder | WHR | SHBG | Sexes combined | Effect of mediator on outcome | -0.10 | 0.04 | 0.012 | -0.09 | 0.03 | 0.009 | -0.10 | 0.06 | 0.067 |
| Bipolar disorder | SI | SHBG | Sexes combined | Intercept | 2.8E-04 | 2.8E-04 | 0.317 | -1.1E-03 | 4.9E-04 | 0.031 | -6.5E-05 | 4.8E-04 | 0.892 |
| Bipolar disorder | SI | SHBG | Sexes combined | Effect of exposure on outcome | 0.01 | 0.01 | 0.109 | 0.03 | 0.01 | 0.009 | 0.01 | 0.01 | 0.149 |
| Bipolar disorder | SI | SHBG | Sexes combined | Effect of mediator on outcome | 0.01 | 0.02 | 0.656 | 0.02 | 0.02 | 0.471 | 0.01 | 0.04 | 0.732 |
| Bipolar disorder | DrnkWk | SHBG | Sexes combined | Intercept | 4.3E-04 | 8.0E-04 | 0.588 | -2.9E-03 | 2.2E-03 | 0.193 | 6.5E-04 | 9.6E-04 | 0.503 |
| Bipolar disorder | DrnkWk | SHBG | Sexes combined | Effect of exposure on outcome | 3.0E-03 | 0.01 | 0.816 | 0.05 | 0.04 | 0.201 | 1.3E-03 | 0.01 | 0.919 |
| Bipolar disorder | DrnkWk | SHBG | Sexes combined | Effect of mediator on outcome | 0.09 | 0.08 | 0.241 | 0.08 | 0.08 | 0.300 | 0.06 | 0.09 | 0.512 |

*CRP = C-reactive protein, SI = smoking initiation, DrnkWk = alcohol drinks per week, AD = alcohol dependence, WHR = waist-hip-ratio*
